# Metabolic stress and disease-stage specific basigin expression of peripheral blood immune cell subsets in COVID-19 patients

**DOI:** 10.1101/2020.09.18.20194175

**Authors:** Peter J. Siska, Katrin Singer, Jana Klitzke, Nathalie Kauer, Sonja-Maria Decking, Christina Bruss, Carina Matos, Kristina Kolodova, Alice Peuker, Gabriele Schönhammer, Johanna Raithel, Dirk Lunz, Bernhard Graf, Florian Geismann, Matthias Lubnow, Matthias Mack, Peter Hau, Christopher Bohr, Ralph Burkhardt, Andre Gessner, Bernd Salzberger, Frank Hanses, Florian Hitzenbichler, Daniel Heudobler, Florian Lüke, Tobias Pukrop, Wolfgang Herr, Daniel Wolff, Hendrik Poeck, Christoph Brochhausen, Petra Hoffmann, Michael Rehli, Marina Kreutz, Kathrin Renner

**Affiliations:** Department of Internal Medicine III, University Hospital Regensburg, 93053 Regensburg, Germany; Department of Otorhinolaryngology, University Hospital Regensburg, 93053 Regensburg; Regensburg Center for Interventional Immunology, University of Regensburg, 93053 Regensburg, Germany; Department of Anesthesiology, University Hospital Regensburg, 93053 Regensburg, Germany; Department of Internal Medicine II University Hospital Regensburg, 93053 Regensburg Germany; Department of Nephrology, University Hospital Regensburg, 93053 Regensburg, Germany; Wilhelm Sander-NeuroOncology Unit and Department of Neurology, University Hospital Regensburg, 93053 Regensburg, Germany; Institute of Clinical Chemistry and Laboratory Medicine, University Hospital Regensburg, 93053 Regensburg, Germany; Institute for Clinical Microbiology and Hygiene, University Hospital Regensburg, 93053 Regensburg, Germany; Department for Infection Control and Infectious Diseases, University Hospital Regensburg, 93053 Regensburg, Germany; Emergency Department, University Hospital Regensburg, 93053 Regensburg, Germany; Bavarian Cancer Research Center, 93053 Regensburg, Germany; Institute of Pathology, University Regensburg, 93053 Regensburg, Germany; Central Biobank Regensburg, University Hospital and University Regensburg, 93053 Regensburg, Germany

**Keywords:** SARS-CoV-2, COVID-19, immune, T cells, monocytes, metabolism, ROS, mitochondria, basigin, CD147

## Abstract

Coronavirus disease 2019 (COVID-19) is driven by dysregulated immune responses yet the role of immunometabolism in COVID-19 pathogenesis remains unclear. By investigating 47 patients with confirmed SARS-CoV-2 infection and 16 uninfected controls, we found an immunometabolic dysregulation specific for patients with progressed disease that was reversible in the recovery phase. Specifically, T cells and monocytes exhibited increased mitochondrial mass, accumulated intracellular ROS and these changes were accompanied by disrupted mitochondrial architecture. Basigin (CD147), but not established markers of T cell activation, was up-regulated on T cells from progressed COVID-19 patients and correlated with ROS accumulation, reflected in the transcriptome. During recovery, basigin and ROS decreased to match the uninfected controls. *In vitro* analyses confirmed the correlation and showed a down-regulation of ROS by dexamethasone treatment. Our findings provide evidence of a basigin-related and reversible immunometabolic dysregulation in COVID-19.

## Introduction

In February 2020 Zhu and colleagues described a novel coronavirus named 2019-nCoV detected in patients with pneumonia of unknown cause in Wuhan, China (Zhu et al., 2020, Huang et al., 2020). Infection with the severe acute respiratory syndrome coronavirus (SARS-CoV-2) causes COVID-19, an illness of varying degrees. The vast majority of COVID-19 cases presents with a mild or a moderately symptomatic infection. However, a subset of individuals progress to develop a severe disease and there is increasing evidence that critical cases are driven by dysregulated host immune responses to SARS-CoV-2 infection allowing virus persistence, causing lung damage and systemic inflammation (De Biasi et al., 2020, Hadjadj et al., 2020, Lee et al., 2020). Infectious diseases change metabolic processes, e.g. in infected cells to support viral replication or in immune cells fighting the infection. Recently Codo and colleagues reported that Sars-CoV-2-infected monocytes up-regulate glycolysis and elevated glucose levels support viral replication and pro-inflammatory cytokine expression (Codo et al., 2020). These data suggest a link between cellular metabolism and systemic host metabolism for disease progression. In line, patients with type 2 diabetes have a greater risk of developing severe disease (Ayres, 2020). Obesity, together with age, is the major risk factor for diabetes, hyperglycemia and dyslipidemia that can result in an imbalance in T cell sub-populations (de Candia et al., 2019). Accordingly, high-fat diet strongly affects T cell metabolism and function, supporting a pro-inflammatory T cell phenotype (Mauro et al., 2017). Interestingly, systemic metabolic changes were observed in COVID-19 patients (Song et al., 2020) and metabolic treatment with cholesterol lowering drugs such as statins can reduce COVID-19 mortality (Zhang et al., 2020).

T cells are central players in adaptive immunity and are essential for controlling viral infections including SARS-CoV-2 (Le Bert et al., 2020). To initiate an immune response, pathogen-specific T cells need to activate and to expand. This is accompanied by an extensive metabolic reprogramming and activated T cells increase expression of nutrient transporters such as glucose transporter GLUT1 and enzymes involved in glycolysis (Geltink et al., 2018). Furthermore, effector T cells contain higher numbers of mitochondria than naive cells (Ron-Harel et al., 2016). Mitochondria produce ATP via oxidative phosphorylation (OXPHOS) and are important for cellular processes such as apoptosis (Ron-Harel et al., 2016) and antiviral defenses (Seth et al., 2005). Besides nutrients, OXPHOS requires oxygen, which is often limiting in patients with COVID-19 (Couzin-Frankel, 2020). Extraordinarily low blood-oxygen levels are often found in infected patients and thus immune cells experience hypoxia and may induce hypoxia inducible factors (HIFs) such as HIF-1a, a transcriptional effector of the hypoxic response. Some studies showed HIF-1a functions as a negative regulator of T cell responses and that O_2_ is required for T cell effector functions (McNamee et al., 2013). To preserve energy production and effector functions, T cells experiencing low O_2_ and glucose levels can enhance fatty acid catabolism as an metabolic escape mechanism (Zhang et al., 2017).

Reduced oxygen saturation and hypoxia lead to generation and accumulation of reactive oxygen species (ROS) by mitochondria (Jezek et al., 2018). Oxidative stress plays an important role in the pathogenesis of viral infections and sepsis, and it has been suggested that COVID-19 pathogenesis is also related to hypoxia and oxidative stress (Laforge et al., 2020). As mitochondrial superoxide leads to an aberrant T cell development, Case et al suggested that manipulations of mitochondrial superoxide levels may significantly alter clinical outcomes of patients with viral infections (Case et al., 2011). Based on this hypothesis, ongoing studies include COVID-19 patients treated with antioxidant drugs such as vitamin C to prevent ROS-induced damage and suppression of antiviral T cells (Cavezzi et al., 2020).

Hypoxia, mitochondrial impairment and the associated generation of ROS have been shown to stabilize CD147 expression (De Saedeleer et al., 2014). CD147, also called basigin or extracellular matrix metalloproteinase inducer (EMMPRIN), is a transmembrane glycoprotein interacting with several extracellular and intracellular partners and can mediate the entry of various viruses (Pushkarsky et al., 2001, Vanarsdall et al., 2018, Watanabe et al., 2010), possibly also for SARS-CoV and SARS-CoV-2 (Chen et al., 2005, Wang et al., 2020). In contrast to angiotensin-converting enzyme 2 (ACE2), the established SARS-CoV receptor (Radzikowska et al., 2020), basigin is broadly expressed in different cell types and tissues (Chen et al., 2005, Muramatsu, 2016). Intriguingly, SARS-CoV proteins do not directly interact with basigin. Instead, the N protein of SARS-CoV binds to cyclophilins (Cyp), which are basigin ligands (Chen et al., 2005). In addition, cyclophilin A (CypA) plays a critical role in the viral replication and Cyp inhibitors such as cyclosporine A (CsA) can block virus replication (Tanaka et al., 2013). Interestingly, CypA is a leukocyte chemoattractant and increased levels of extracellular CypA have been documented in inflammatory diseases (Bukrinsky, 2015). Given the roles of basigin and Cyp in viral diseases, a clinical trial investigated the impact of basigin blocking by Meplazumab, a humanized anti-CD147 antibody and reported improved outcome of patients with COVID-19-associated pneumonia (Bian et al., 2020). Basigin is weakly expressed on naïve T cells but up-regulated on activated cells. Based on this finding, ABX-CBL, an immunoglobulin M murine monoclonal anti-CD147 antibody was successfully used in a trial with patients with steroid-refractory acute graft-versus-host disease (Deeg et al., 2001). Apart from its role for virus entry, basigin tightly associates with GLUT1, the amino acid transporter CD98, the lactate transporters (monocarboxylate transporter/MCT 1, 3 and 4) and CD44 and is recognized by lectins, such as galectin-3 and E-selectin (Muramatsu, 2016). These molecular interactions explain the central role of basigin in energy metabolism, cell motility and activation. However, basigin deficient mice are more active in mixed lymphocyte reactions, indicating the suppressive role of basigin on T cell activation which might be related to the inhibition of nuclear factor of activated (NFATs) in T cells (Hahn et al., 2015). In addition, basigin suppresses NOD2, an important innate immunity component (Muramatsu, 2016).

Metabolic features of immune cells strongly determine the outcomes of an immune response and a variety of publications reported dysregulated innate and adaptive immune pathways in COVID-19 patients.

Therefore, we studied immune cell metabolism of COVID-19 patient subgroups with different degrees of disease severity with or without pre-existing metabolic comorbidities. Our data show T cell metabolic changes in mitochondrial mass, architecture and ROS production as well as in fatty acid uptake in the course of disease that are (1) not related to classical activation markers, (2) stage specific, (3) basigin-related and (4) reversed in recovering patients to reach levels of healthy controls. Our data suggest that immunometabolic dysregulation contributes to the dysfunctional T cell response in COVID-19 patients.

## Results

### Altered T cell and NK cell distribution in COVID-19 patients

COVID-19 has previously been associated with changes in the peripheral blood immune compartment (Lucas et al., 2020, Zhou et al., 2020, Qin et al., 2020). Here we analyzed peripheral blood immune cell populations from age-matched non-infected controls ("control" group), asymptomatic and mildly symptomatic SARS-CoV-2 positive subjects ("no symptoms/mild" group), moderately ill ("moderate"), severely to critically ill patients ("severe/critical" group) and patients recovering from a severe/critical COVID-19 ("recovering" group). Classification details and patient characteristics can be found in the methods section and in Table 1. Blood counts revealed no significant changes in immune cell frequencies in COVID-19 patients with progressed disease even though lymphocytes were decreased by trend (**Table 1**).

**Table 1.** Clinical characteristics of COVID-19 patients. a Laboratory values from the day of subsequent single-cell analyses b Statistical tests used: Kruskal-Wallis (age), chi-squared test after Pearson (all categorical variables), students t-test (ferritin), one-way ANOVA (leukocytes and all other laboratory values). For continuous variables, data are medians and interquartile range. For categorical variables, data counts and percentages of patients where data were available.

|  | None/mild (n=15) | Moderate (n=5) | Severe/critical (n=27) | p-value<sup>b</sup> |
| --- | --- | --- | --- | --- |
| **Clinical characteristics** |  |  |  |  |
| Age (y) | 57.9 (43.9-67.2) | 60.2 (49.8-74.8) | 58.2 (46.7-68.1) | 0.726 |
| Male gender | 7/15 (47%) | 3/5 (60%) | 17/27 (63%) | 0.588 |
| Prior malignancy | 5/13 (38%) | 1/5 (20%) | 5/27 (19%) | 0.377 |
| Prior transplantation and/or immunosuppression | 1/13 (8%) | 2/5 (40%) | 2/27 (7%) | 0.093 |
| Prior type 2 diabetes and/or adipositas | 5/14 (36%) | 1/5 (20%) | 10/27 (37%) | 0.761 |
| Prior hypertension | 5/14 (36%) | 2/4 (50%) | 11/24 (46%) | 0.749 |
| Prior arrhythmia | 2/14 (14%) | 1/4 (25%) | 7/27 (26%) | 0.690 |
| Prior cardiovascular comorbidity | 7/14 (50%) | 4/5 (80%) | 13/27 (48%) | 0.416 |
| Mechanical ventilation | 0/14 (0%) | 0/4 (0%) | 24/27 (89%) | <0.001 |
| Extracorporeal membrane oxygenation | 0/14 (0%) | 0/4 (0%) | 11/27 (41%) | <0.01 |
| Antibody response | 3/6 (50%) | 2/3 (67%) | 22/22 (100%) | <0.01 |
| Death | 0/14 (0%) | 0/5 (0%) | 4/27 (15%) | 0.214 |
| **Laboratory values<sup>a</sup>** |  |  |  |  |
| Ferritin (ng/ml) | 402.5 (247-3279) | na | 1461 (863-3705) | 0.4072 |
| Leukocytes ( /nl) | 6.1 (3.8-9.4) | 4.89 (4.2-6.6) | 8.05 (5.1-16.5) | 0.1642 |
| Lactate dehydrogenase (U/l) | 204.5 (152.5-215.3) | 379 (251-387) | 284 (247-381.5) | 0.1147 |
| C-reactive protein (mg/l) | 13.7 (5.5-30.3) | 100 (33.9-130) | 59.8 (19.8-97.8) | 0.0487 |
| Neutrophils (%) | 62.7 (51.6-68.1) | 65.9 (34.2-85.4) | 70.55 (59.6-81.6) | 0.5520 |
| Basophiles (%) | 0.4 (0.2-0.4) | 0.2 (0.2-1.4) | 0.55 (0.2-0.9) | 0.5999 |
| Eosinophils (%) | 2.5 (1.5-3.4) | 1.8 (0.5-2) | 3 (0.6-5.0) | 0.6176 |
| Monocytes (%) | 8.3 (8-9.3) | 8.8 (7.6-18.3) | 8.1 (5.8-10.5) | 0.2499 |
| Lymphocytes (%) | 26.8 (20.2-3) | 21.9 (6.3-45.5) | 17.3 (11.3-24.1) | 0.6218 |
| Immature granulocytes (%) | 0.3 (0.2-0.5) | 2.2 (0.8-3.4) | 1.2 (0.5-3.4) | 0.2006 |

Fresh samples were not subject to any manipulation such as mononuclear cell enrichment or cryopreservation. Multi-parameter flow cytometry was performed to discriminate cells based on expression of CD14/CD11b (monocytes) and CD16+/-(monocyte subsets), CD66b/CD11b (granulocytes), CD56 (NK cells), CD3 (T cells), T cell subsets expressing CD4, CD8 and FOXP3, CD19 (B cells) and CD19/CD38/IgA/IgM/IgG (plasmablasts) (**Figure 1A**). The frequencies of monocytes, including CD16+ and CD16-monocyte subpopulations (**Figures 1B and S1A**), granulocytes (**Figure 1C**) and FOXP3+ CD4+ T cells (**Figure 1D**) were not significantly altered in COVID-19 patients. However, patients in the severe/critical group showed low NK cell and T cell frequencies and these changes persisted in recovering patients (**Figure 1E,F**). While a decrease in T cells has been described for patients with progressed SARS-CoV and SARS-CoV-2 infections (Lucas et al., 2020, Li et al., 2004), we observed lower frequencies of T cells already in the group of COVID-19 patients with moderate symptoms (**Figure 1F**). Notably, patients with fatal disease showed the lowest number of T cells and NK cells. Further analyzing T cell subsets, similar CD8+ and CD4+ T cell frequencies were observed in the severe/critical group, as compared to control subjects. However, patients from the moderate group presented a shift towards CD8+ T cells (**Figure 1G**). To study the dynamics of T cell changes in the severe/critical group, we clustered paired samples according to the time after the onset of symptoms. While CD8+ T cells increased and CD4+ T cells decreased during the first phase of infection in most patients, an opposite trend was observed during the second phase (**Figure S1B**). Lastly, studying the peripheral blood B cell compartment, we observed a prominent increase both in bulk (**Figure 1H**) as well as in IgA+ and IgG+, but not IgM+ plasmablasts in SARS-CoV-2 infected subjects (**Figure 1I**). In addition, plasmablast formation, marked by decreased frequency of CD20+ B cells, was observed in critically ill COVID-19 patients during the hospital stay (**Figure S1C**).

**Figure 1.**
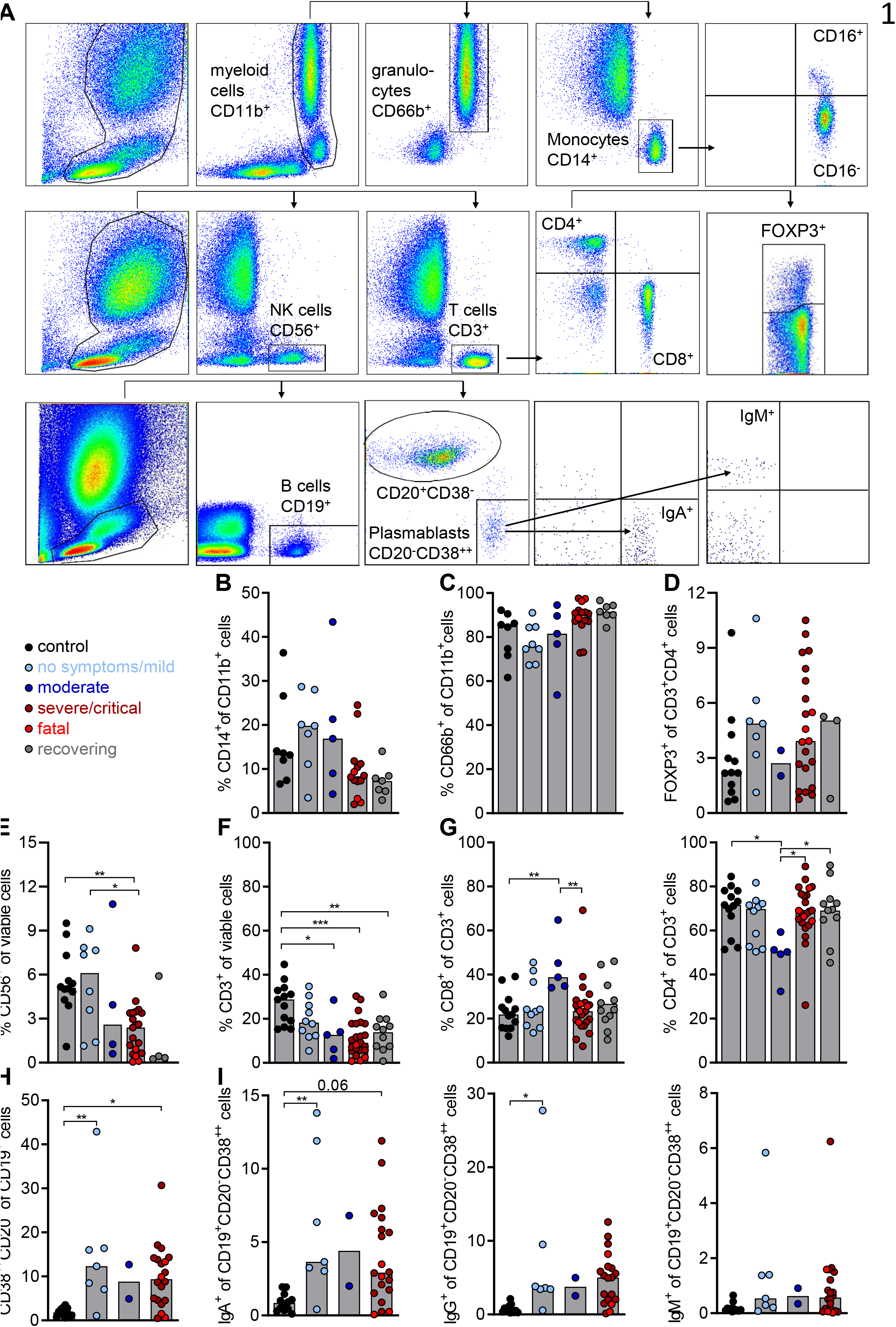
Altered T cell and NK cell distribution in COVID-19 patients. Blood of donors was drawn and processed on the same day. Erythrocytes were removed by ACK lysis and immune cell populations were determined by surface marker staining and subsequent analysis by flow cytometry. (**A**) Representative plots displaying the gating strategy for specific immune cell populations. CD14+ (**B**) and CD66+ (**C**) cells among CD11b+ myeloid cells are shown in specified COVID-19 patient subgroups and healthy controls. (**D**) The frequency of regulatory T cells was determined by intracellular FOXP3 staining in CD3+CD4+ T cells. Percentage of NK cells (**E**), CD3+ T cells (**F**), and CD8+ and CD4+ T cell subpopulations (**G**), is displayed. (**H**) Percentage of B cell blasts among CD19+ B cells, and frequencies of IgA+, IgG+ and IgM+ B cell blasts (**I)**, were determined. Shown is the median, each symbol represents one donor (one-way ANOVA, Bonferroni multiple comparisons test, *p<0.05, **p<0.01, ***p<0.001).

Collectively, altered NK and T cell frequencies, skewed T cell subset distribution and plasmablast induction were observed in COVID-19 patients.

### T cells from progressed COVID-19 patients show high expression of basigin, but are not infected with SARS-CoV-2

In addition to immune cell sub-population frequencies, the activation state might determine the clinical outcome of viral diseases. Therefore, the activation and differentiation state of T cells and monocytes was assessed in COVID-19 patients and control subjects. While CD8+ T cells but not CD4+ T cells from patients in the severe/critical group showed a trend towards increased CD25 expression (**Figure 2A**), no differences were observed for PD-1 expression. Nevertheless, patients with fatal disease showed a trend to cluster in PD-1-high T cell subpopulations (**Figure 2B**). No differences were detected in CCR7 expression in both T cell subsets (**Figure 2C**). Studying HLA-DR expression on monocytes, we observed a trend towards a decrease in the severe/critical group and all deceased patients clustered in a severe/critical subgroup with low monocyte HLA-DR expression (**Figure S2A**). No differences were observed when studying HLA-DR expression of CD16+ and CD16-monocyte subpopulations (**Figure S2B**).

**Figure 2.**
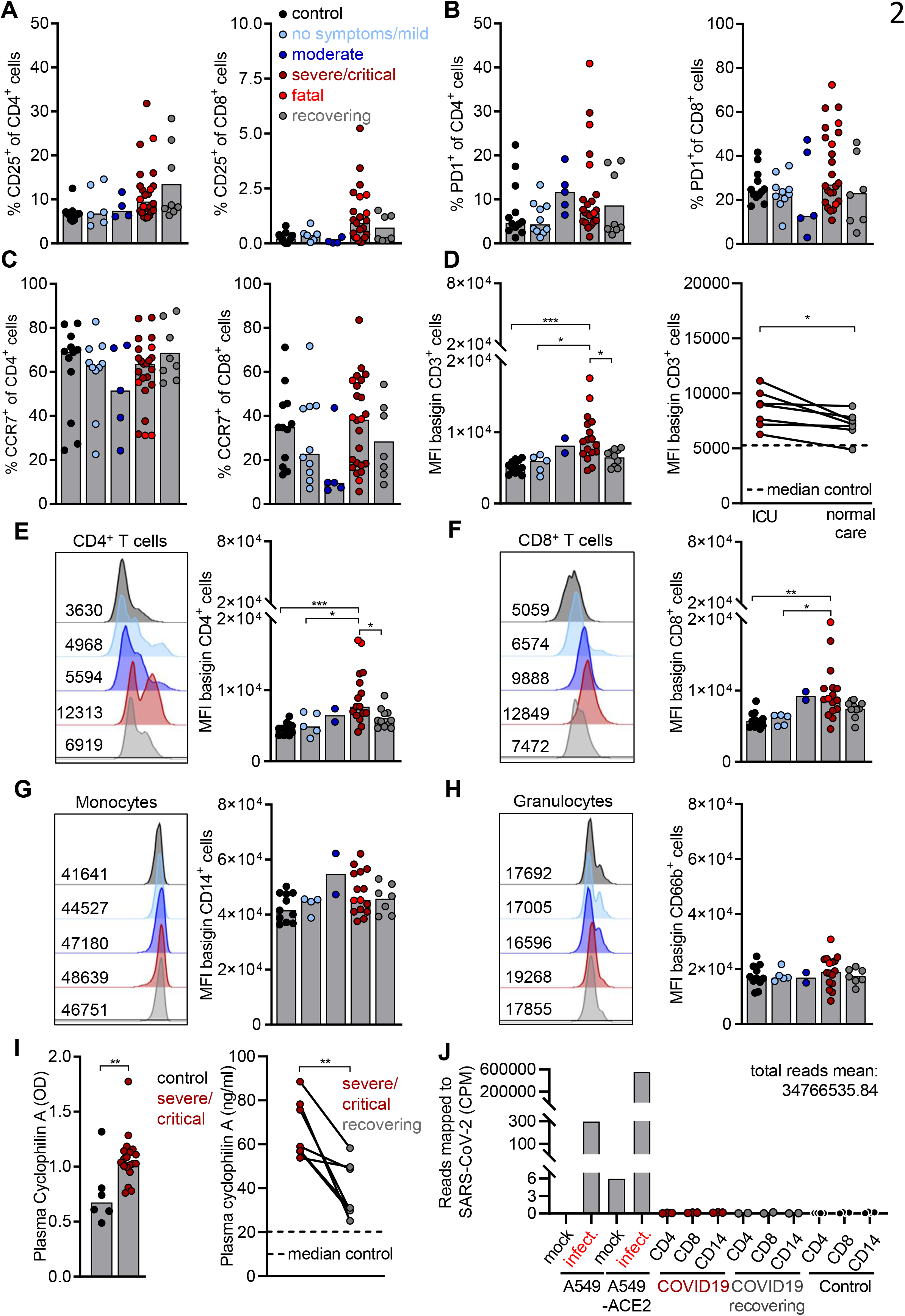
T cells from progressed COVID-19 patients show high expression of basigin, but are not infected with SARS-CoV-2. **(A-H)** Peripheral blood was drawn and processed on the same day. Erythrocytes were removed by ACK lysis and immune cell populations were stained for population-specific surface markers. CD25 (**A**), PD-1 (**B**) and, CCR7 (**C**) expression on CD4+ and CD8+ T cell subpopulations was analyzed. Shown is the median and each symbol represents one donor. Basigin expression was determined on CD3+ T cells in all groups (left) and paired samples (right) of patients taken at the intensive care unit (ICU) and after returning to normal care unit (normal care) (**D**). Shown is the median of all groups, each symbol represents one donor (one-way ANOVA, Bonferroni multiple comparisons test, *p<0.05, ***p<0.001), line represents a paired sample (paired Student’s t-test). (**E-H**) Basigin expression on CD4+ (**E**) and CD8+ (**F**) subpopulations as well as on monocytes (**G**) and granulocytes (**H**). Shown is a representative blot of each group and summarized data are displayed as median levels with a single symbol for each donor (one-way ANOVA, Bonferroni multiple comparisons test, *p<0.05, **p<0.01 ***p<0.001). (**I**) Cyclophilin A levels were determined in the plasma of controls and COVID-19 patients by ELISA. Shown are median levels, each symbol represents one donor (unpaired Student’s t-test), and paired samples of patients taken at the intensive care unit (ICU) and after returning to normal care unit (normal care, paired Student’s t-test). (**J**) CD4+ T cells, CD8+ T cells and CD14+ monocytes were sorted from blood of four healthy controls and COVID-19 patients at the ICU (n=3) or in recovery (n=2, matched samples). RNA sequencing was performed and reads were mapped to the viral (SARS-CoV-2, NC_045512.2) genome. Published data of SARS-CoV-2 infected A549 cells was identically processed as a control (Blanco-Melo et al., 2020). Shown are counts per million of reads mapped to the SARS-CoV-2 genome.

CD147 (basigin, also known as EMMPRIN) has been previously suggested as a marker of T cell activation but also as a negative regulator of T cell proliferation (Kasinrerk et al., 1992, Supper et al., 2016, Hahn et al., 2015, Hu et al., 2010). As opposed to PD-1, CD25 and CCR7, both bulk T cells (**Figure 2D**) and CD4+ and CD8+ T cell subtypes (**Figure 2E,F**) showed an increased expression of basigin in patients with progressed COVID-19. Interestingly, while altered T cell frequency persisted in recovering patients (**Figure 1F and S2C)**, T cell basigin expression decreased in patients during recovery from COVID-19 to levels comparable with uninfected control subjects (**Figures 2D-F and S2D,E**). Monocytes showed high basal basigin expression but in contrast to T cells, the levels were similar among the studied groups (**Figure 2G**). Similarly, no differences in basigin expression were observed in granulocytes (**Figure 2H**). In concert with cyclophilin (Pawlotsky, 2020), basigin can mediate the entry of human coronaviruses (Chen et al., 2005). Accordingly, cyclophilin targeting drugs can inhibit coronavirus replication (Carbajo-Lozoya et al., 2014, Tanaka et al., 2013) and an anti-basigin antibody is being evaluated for COVID-19 therapy (Bian et al., 2020). Therefore, we measured plasma cyclophilin A and observed elevated levels in the severe/critical group as compared to control samples, while recovering patients showed a decrease in cyclophilin concentrations (**Figure 2I**). We next assessed, whether immune cells might have been infected with SARS-CoV-2. Published data from A549 cells infected with SARS-CoV-2 (Blanco-Melo et al., 2020) indicated a high amount of detectable viral RNA, which further increased if cells were transfected to express angiotensin-converting enzyme 2 (ACE2). Cell sorting and subsequent RNA sequencing however revealed no evidence for SARS-CoV-2 infection of CD4+ and CD8+ T cells or CD14+ monocytes of critically ill COVID-19 patients or healthy donors (**Figure 2J**).

Collectively, basigin, but not the established markers of T cell activation and differentiation, was strongly up-regulated on T cells from progressed COVID-19 patients and decreased during recovery. Despite increased basigin expression and high cyclophilin A plasma levels, immune cells from COVID-19 patients were likely not infected with SARS-CoV-2.

### Glucose and fatty acid uptake by immune cells distinguishes COVID-19 patient subgroups

Increasing evidence points to basigin as a regulator of cellular metabolism as it tightly associates with monocarboxylate transporters (MCTs), transporters for glucose (GLUT) and amino acids (CD98), likely coordinating glucose and amino acids metabolism (Muramatsu, 2016). In addition, basigin signaling can inhibit fatty acid metabolism via modulation of PPARa (Li et al., 2015). Therefore, the capacity of T cells and myeloid cells from COVID-19 patients to take up glucose, fatty acids and the expression of the amino acid transporter CD98 was assessed next in fresh patient samples.

The glucose uptake capacity of CD4+ and CD8+ T cells as well as of CD11b+ myeloid cells did not differ among the studied patient groups (**Figure 3A**). Similarly, T cell expression of CD98 was similar among the patient groups (**Figure S3A**). T cell glucose uptake capacity was highly variable between COVID-19 patients and we asked whether patient pre-existing comorbidities contribute. Diabetes and obesity are associated with unfavorable COVID-19 prognosis (Ayres, 2020) and we observed decreased CD3+ T cell, but not myeloid cell counts in patients with pre-existing diabetes and/or obesity (**Figure 3B**). Interestingly, these patients also showed a decreased glucose uptake capacity of T cells and myeloid cells compared to controls (**Figure 3C**), while a separation of patients according to cardiovascular disease or age did not show significant differences in immune cell glucose uptake (**Figures 3D and S3B**). Immune cells of diabetic COVID-19 patients might compensate low glucose uptake with increased fatty acid metabolism. However, no differences in fatty acid uptake were detected in patients with diabetes (**Figure S3C**). Interestingly, regardless of comorbidities, we observed an increased fatty acid uptake in T cells from patients with no or mild symptoms (**Figure 3E**). Although T cell fatty acid uptake in the severe/critical group was low, a clustering of paired samples according to the time after the onset of symptoms revealed an increase of T cell fatty acid uptake between day 0 and 40 (**Figure S3D**). While myeloid cells did not show an increase in the none/mild group, patients from the severe/critical group presented a strong decrease in fatty acid uptake, with deceased patients being the lowest (**Figure 3F**). Patients from the none/mild group showed increased T cell fatty acid uptake and we next assessed the dynamics of this metabolic phenotype after complete disease resolution. Fatty acid uptake of CD4+ and CD8+ T cells as well as of their respective subsets was analyzed in three COVID-19 patients from one family with mild symptoms over the course of 7 weeks after the onset of symptoms (**Figure 3G**). Interestingly, the capacity of T cells to take up fatty acids decreased over time. Notably, this phenomenon occurred 4 weeks after the onset of symptoms, and was observed in bulk CD8+ and CD4+ T cells as well as in major T cell subpopulations (**Figures 3H and S3E**).

**Figure 3.**
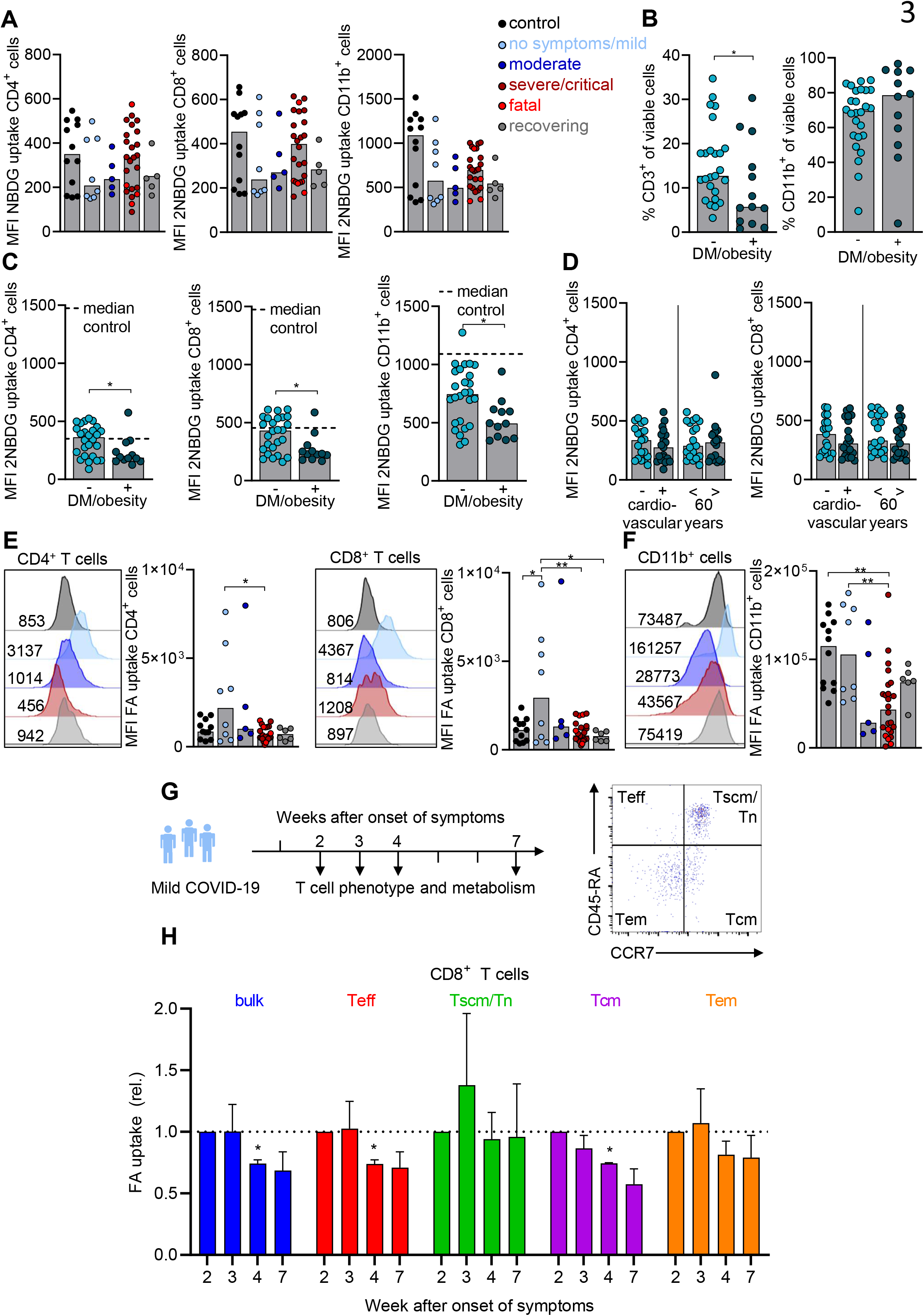
Glucose and fatty acid uptake by immune cells distinguishes COVID-19 patient subgroups. Blood of donors was drawn and processed the same day. Erythrocytes were removed by ACK lysis and immune cell populations were stained for population-specific surface markers. (**A**) Determination of 2NBDG uptake by flow cytometry as measurement of glucose metabolism in CD4+ and CD8+ T cells and CD11b+ myeloid cells. Shown is the median, each symbol represents one donor. (**B**) Frequencies of CD3+ T cells and CD11b+ myeloid cells compared in diabetic/obese versus non-diabetic/obese patients. Shown is the median, each symbol represents one patient (Mann-Whitney U test, *p<0.05). (**C**) Comparison of 2NBDG uptake in CD4+ and CD8+ T cells and CD11b+ myeloid cells in diabetic and/or obese versus non-diabetic and/or obese patients. Shown is the median, each symbol represents one patient (Mann-Whitney U test, *p<0.05). (**D**) 2NBDG uptake in CD4+ and CD8+ T cells from COVID-19 patients with or without depicted pre-existing comorbidity. (**E-F**) Determination of BODIPY 500/510 C1, C12 uptake by flow cytomtery as a measurement of fatty acid (FA) uptake in CD4+ and CD8+ T cells (**E**) and CD11b+ myeloid cells (**F**). Shown is a representative blot of each group and summarized data are displayed as median levels, a single symbol represents one donor (one-way ANOVA, Bonferroni multiple comparisons test, *p<0.05, **p<0.01). (**G**) Sampling scheme of blood and gating strategy based on CD45RA and CCR7 surface staining of T cell subsets from 3 patients of one family with mild COVID-19 for the analysis of fatty acid uptake in CD8+ T cells (**H**). Shown is the mean and SEM of values normalized to the first blood sample withdrawn (Mixed-effects model with the Geisser-Greenhouse correction, with Bonferroni’s multiple comparisons test, *p<0.05).

Taken together, immune cells from COVID-19 patient subgroups were characterized by different capacities to take up nutrients. While metabolic comorbidities affected immune cell glucose uptake capacity, increased T cell fatty acid uptake was a hallmark of COVID-19 patients with no or mild symptoms and decreased during convalescence.

### ROS accumulation in T cells from progressed COVID-19 patients is reversible and correlates with disrupted mitochondrial architecture and expression of ROS-inducible and basigin-related genes

T cells from patients with viral or malignant diseases can enter a state of metabolic exhaustion with decreased nutrient uptake, accompanied by accumulation of reactive oxygen species (ROS) (Siska et al., 2017, Bengsch et al., 2016). We observed low nutrient uptake in freshly analyzed immune cells from critically ill COVID-19 patients, suggesting a similar immunometabolic state. While basigin has been associated with increased cellular metabolic activity (Muramatsu, 2016), metabolic stress and reactive oxygen species (ROS) can also induce and stabilize basigin expression (De Saedeleer et al., 2014). In line, we observed a correlation of T cell basigin expression with levels of intracellular ROS in COVID-19 patients (**Figure 4A**). Moreover, patients in the severe/critical group showed high T cell ROS levels that significantly decreased in convalescence, suggesting an immunometabolic recovery (**Figure 4B**). While high basal levels of ROS and basigin were observed in CD14+ myeloid cells, these parameters did not correlate (**Figure 4C**) and myeloid cell ROS levels were not significantly elevated in patients in the severe/critical group (**Figure 4D**). Similarly, ROS levels in neutrophils were comparable between all patient groups analyzed (**Figure 4E**). Dysregulation of cellular redox homeostasis is a hallmark of mitochondrial dysfunction (Jezek et al., 2018). Analyses of the mitochondrial content of T cells and myeloid cells from COVID-19 patients revealed a mitochondrial mass increase in the severe/critical group in T cells and monocytes (**Figure 4F,G**). In contrast to glucose uptake, neither ROS nor the mitochondrial content of T cells were related to a pre-existing comorbidity (**Figure S4A,B**). To correlate the observed metabolic phenomena with mitochondrial morphology, electron microscopy of peripheral blood mononuclear cells from critically ill COVID-19 patients was performed. In line with the observed increase in mitochondrial mass and ROS accumulation, mitochondria of lymphoid cells from COVID-19 patients were enlarged and showed a disrupted cristae architecture as compared to control lymphocytes (**Figure 4H**).

**Figure 4.**
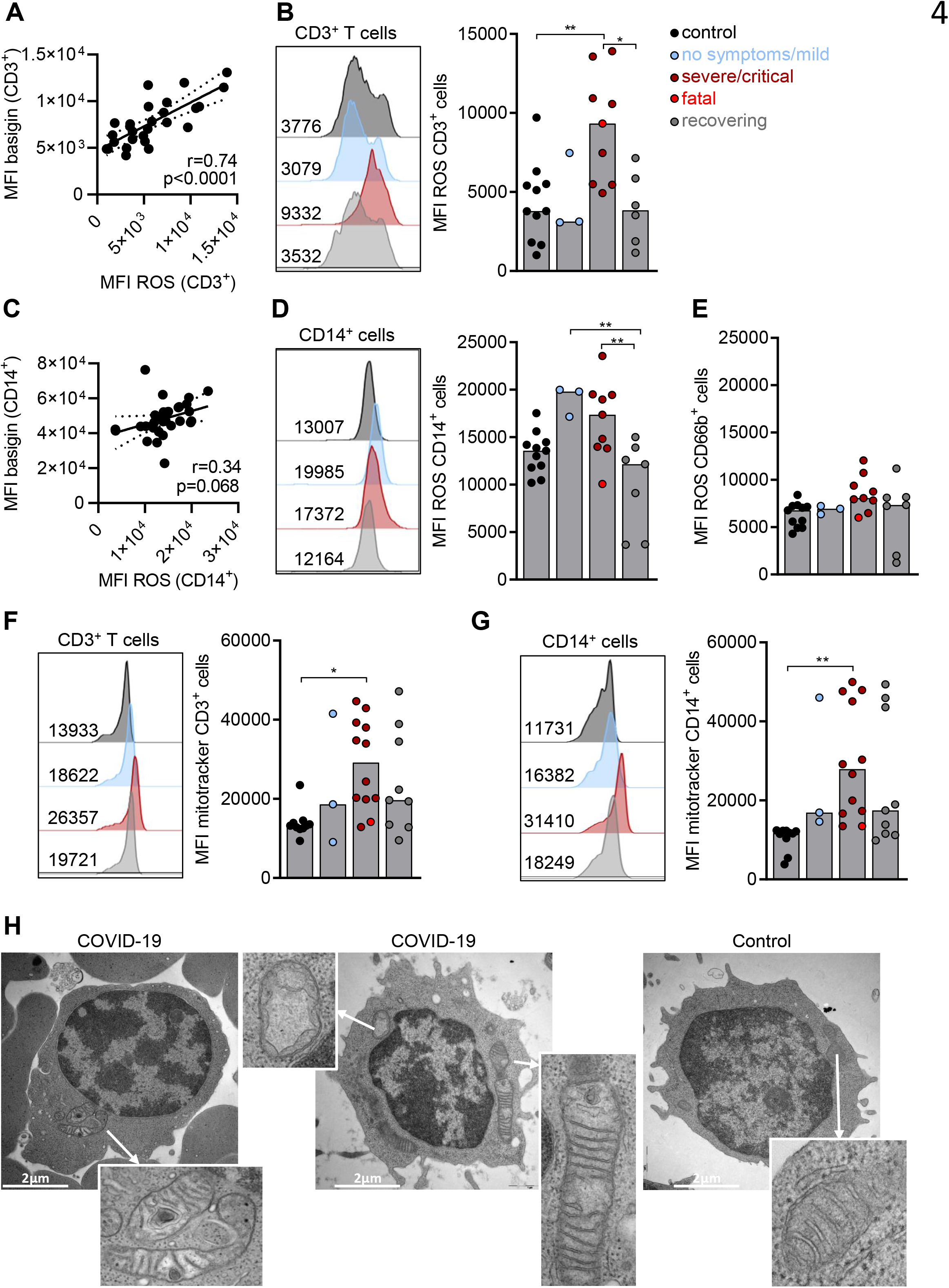
ROS accumulation in T cells from progressed COVID-19 patients is reversible and correlates with high basigin expression and a disrupted mitochondrial architecture. Blood of donors was drawn and processed on the same day. Erythrocytes were removed by ACK lysis and immune cell populations were stained for population-specific surface markers. (**A-E**) Cytosolic ROS levels were determined by DCFDA staining and analyzed by flow cytometry. ROS levels in CD3+ T cells (**A**) correlated to basigin expression (each dot represents a donor, Spearman r and p values are displayed) and displayed for all groups (**B**). Shown is a representative blot of each group and summarized data displayed as median levels and a single symbol for each donor (one-way ANOVA, Bonferroni multiple comparisons test, *p<0.05, **p<0.01). (**C,D**) Correlation of basigin expression and ROS levels in CD14+ monocytes, as in (A,B) and CD66b+ neutrophils (**E**) of all sample groups. (**F,G)** Determination of mitochondrial content by mitotracker staining analyzed by flow cytometry in CD3+ T cells (**F**) and CD14+ monocytes (**G**). Shown is a representative blot of each group and summarized data displayed as median levels and a single symbol for each donor (one-way ANOVA, Bonferroni multiple comparisons test, *p<0.05, **p<0.01). (**H**) Analysis of mitochondrial structure by electron microscopy in lymphocytes of two COVID-19 patients and healthy control lymphocytes. Shown are representative examples.

To validate the observations from the single cell studies, RNA sequencing of T cells and monocytes from COVID-19 patients (three severe/critical patients; two recovering patients) and healthy controls (four individuals) was performed. Samples were enriched for mononuclear cells and cryopreserved prior to flow cytometry sorting. CD4+ T cells from COVID-19 patients from the severe/critical group showed significantly increased expression of 49 genes, while 9 genes were down-regulated (**Figure 5A**). Besides genes associated with immune signaling and function such as GAB2, BCL3 and SOCS3, the expression of SLC16A1 (MCT1), which closely associates with basigin, was increased in CD4+ T cells from COVID-19 patients. Interestingly, CD4+ T cells from COVID-19 patients showed increased expression of furin, that was recently reported to activate SARS-CoV-2 (Hoffmann et al., 2020) (**Figure 5B**). Utilizing the gene set enrichment approach (Liberzon et al., 2015), CD4+ T cells from COVID-19 patients showed a significant enrichment of genes associated with ROS, genes coding for basigin interaction partners and genes related to TNF signaling (**Figures 5C and S5**). CD14+ monocytes showed a COVID-19 specific up-regulation of 174 genes (**Figure 5D**), yet the expression of SLC16A1, furin and several other CD4+ T cell specific genes was not increased. However, genes involved in metabolism such as PPARG or GLUL were increased as well as HIF1A, suggestive for a response to hypoxia (**Figure 5E**). Indeed, transcripts of hypoxia related genes were strongly enriched in monocytes from COVID-19 patients in addition to genes related to TNF signaling (**Figure 5F**).

**Figure 5.**
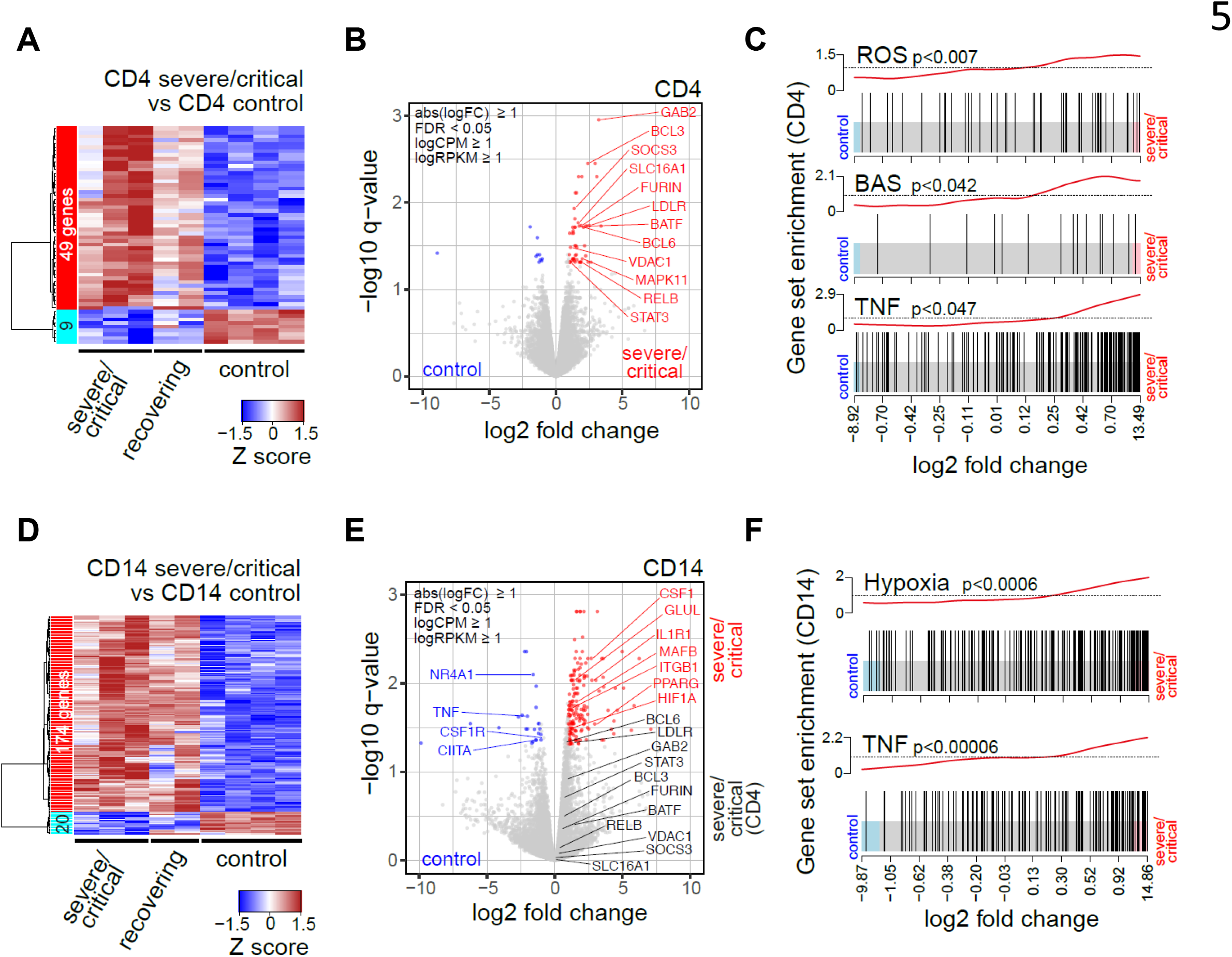
Transcriptional profiling reveals an enrichment of ROS-related genes in CD4+ T cells but not in CD14+ monocytes from COVID-19 patients. Blood samples from COVID-19 patients (n=5, with 3 at the ICU and 2 in recovery) and four healthy controls were taken and mononuclear cells were cryopreserved. Cells were sorted and RNA of immune subpopulations was extracted and subjected to stranded total RNA sequencing. (**A**) Heatmap presenting hierarchically clustered and scaled expression data of sorted CD4+ T cells. Shown are differentially expressed genes in severe/critical patients versus controls filtered for absolute logFC > 1, logCPM and logRPKM > 1 and a FDR < 0.05. Each column corresponds to an individual patient (either severe/critical or recovering) or healthy control. The gene list and expression values can be found in the manuscript supplement. (**B**) Volcano plots of significantly up- or down-regulated genes (red or blue, respectively) in severe/critical patients in sorted CD4+ T cells. Genes with known function in transcriptional or metabolic processes are highlighted. Red, up-regulated, blue, down-regulated in severe/critical patients. (**C**) Gene set enrichment analysis of candidate HALLMARK pathways in CD4+ T cells. (**D**) CD14+ monocytes as in (A). The gene list and expression values can be found in the manuscript supplement. (**E**), as in (B), with additional genes upregulated in CD4+ T cells but not in CD14+ monocytes in black. (**F**) Gene set enrichment analysis of candidate HALLMARK pathways in CD14+ monocytes. HALLMARK pathways include REACTIVE_OXYGEN_SPECIES_PATHWAY (ROS), TNFA_SIGNALING_VIA_NFKB (TNF) and HYPOXIA as well as CD147/basigin interacting proteins (BAS), as defined by STRING analysis (see **Figure S5**).

Collectively, in addition to low nutrient uptake, T cells from progressed COVID-19 patients showed a metabolic dysregulation with increased mitochondrial mass, altered mitochondrial morphology and accumulation of ROS. Transcriptome analyses showed an increased T cell expression of ROS-related genes and basigin interaction partners and suggested a response to hypoxia by monocytes.

### T cell receptor stimulation induced ROS accumulation in COVID-19 basigin-high T cells is reversible through dexamethasone treatment

We observed an association of basigin expression and accumulation of intracellular ROS in T cells from COVID-19 patients in the severe/critical group. To assess, whether external factors contribute, we cultured mononuclear cells from COVID-19 patients and healthy controls *in vitro* in medium containing the antioxidant 2-mercaptoethanol. In addition, cultured cells were treated with anti-CD3/anti-CD28 coated beads to mimic an *in vivo* T cell receptor activation. While uncultured COVID-19 T cells showed elevated ROS and basigin levels, a six-day *in vitro* culture without stimulation lead to a normalization as compared to control cells. Furthermore, activation strongly induced ROS and basigin in T cells and both parameters correlated at the single cell level (**Figures 6A and S6A**).

**Figure 6.**
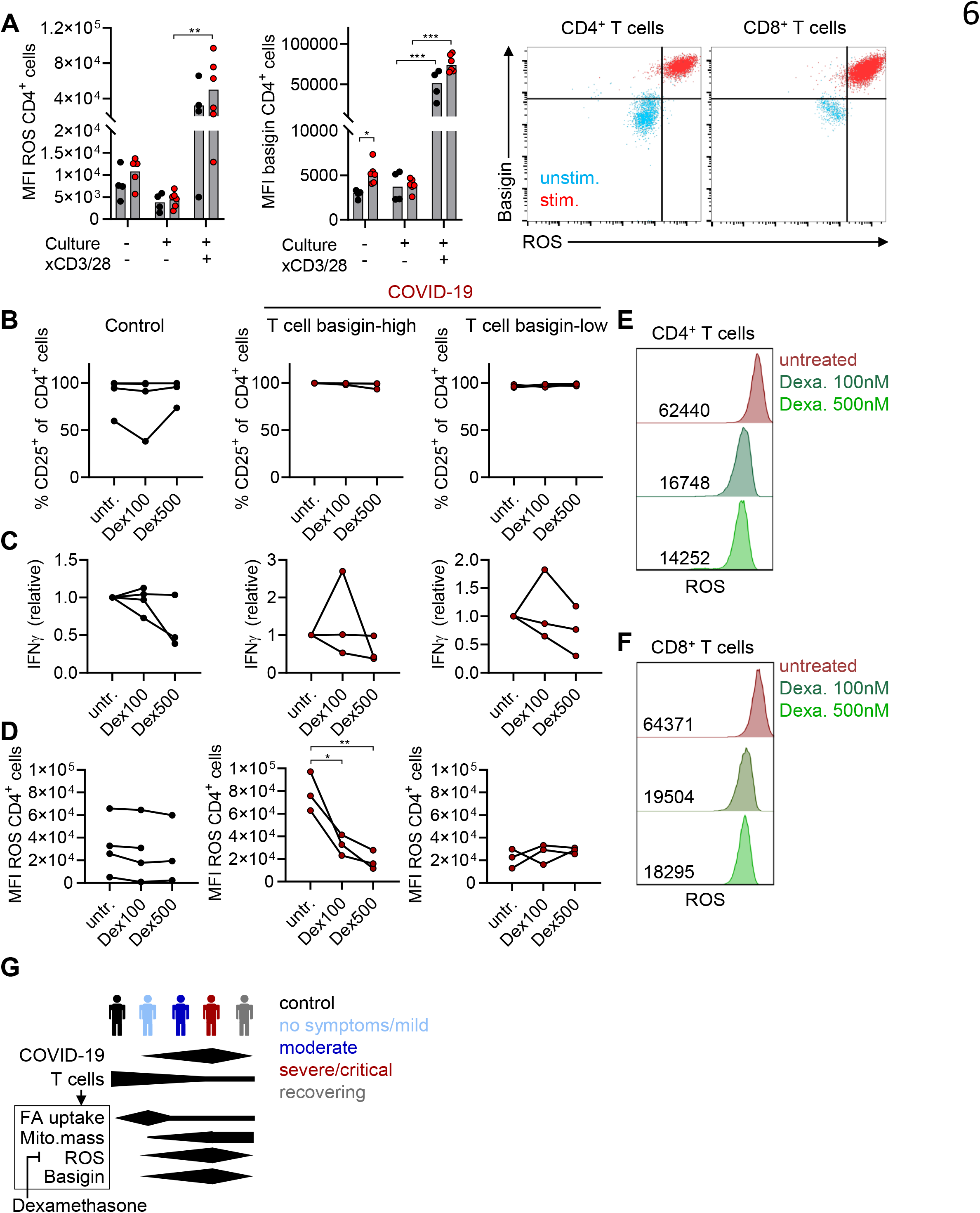
T cell receptor stimulation induced ROS accumulation in COVID-19 basigin-high T cells is reversible through dexamethasone treatment. Mononuclear cells were isolated from controls or COVID-19 patients from the severe/critical group and cryopreserved until analysis. (**A**) ROS and basigin levels were determined in CD4+ T cells of controls and severe/critically ill patients before and after a 6 days culture in medium containing 2-mercaptoethanol with or without anti-CD3/anti-CD28 stimulation. Shown are median levels and a single symbol for each donor (paired Student’s t-test, **p<0.01, ***p<0.001). Representative FACS plots also include CD8+ T cells. (**B-F**) Mononuclear cells of controls and COVID-19 patients with high and low basal basigin expression (as pre-defined in **Figure S6B**) were anti-CD3/anti-CD28 stimulated for six days in the presence or absence of increasing concentrations of dexamethasone (100 nM, 500 nM) and percentage of CD25+CD4+ T cells (**B**), IFN**y** release normalized to untreated cells, measured by ELISA (**C**) and cytosolic ROS levels, determined by DCFDA staining (**D**), were assessed. Shown are paired samples from each donor, represented by a symbol (one-way ANOVA, Bonferroni multiple comparisons test, *p<0.05, **p<0.01). (**E,F**) Representative plots showing the impact of dexamethasone on ROS levels in CD4+ and CD8+ T cells of on critically ill COVID-19 patient with high basal basigin expression.

Therapeutic approaches for COVID-19 are still limited. Nevertheless, recent data suggest that dexamethasone treatment improves the survival of critically ill COVID-19 patients (Group et al., 2020). Dexamethasone is a potent modulator of immune cell responses and lymphocyte metabolism (Eberhart et al., 2011, Serrano et al., 1993) and we tested, whether *in vitro* dexamethasone treatment can revert the T cell metabolic phenotype hallmarked by high basigin expression. *Ex vivo* T cell basigin expression varied strongly (**Figure 2D**) and we next tested, how COVID-19 T cells with high or low basal basigin levels (**Figure S6B**) respond to dexamethasone. While dexamethasone did not affect T cell activation in terms of CD25 expression (**Figures 6B and S6C**) or interferon-y production (**Figure 6C**), it prevented intracellular ROS accumulation in T cells from COVID-19 patients with high basal basigin expression, but not from control subjects or patients with normal T cell basigin levels (**Figures 6D-F and S6D**).

Thus, T cell receptor stimulation induced ROS accumulation, accompanied by basigin up-regulation in T cells from COVID-19 patients and controls. Treatment with dexamethasone decreased T cell ROS accumulation specifically in T cells from patients with high basal basigin levels. Collectively, progressed COVID-19 is associated with increased T cell basigin expression, and a metabolic dysregulation hallmarked by ROS accumulation that is reversed during recovery or through pharmacologic modulation *in vitro* (**Figure 6G**).

## Discussion

It is now clear, that the role of cellular metabolic pathways goes beyond energy production. Intriguingly, immune cell metabolism can steer immune cell function and thereby affect the outcome of inflammation (O’Neill et al., 2016). Here we studied the composition, phenotype and metabolism of immune cells in COVID-19 patients and describe a T cell metabolic phenotype with high accumulation of reactive oxygen species (ROS), elevated mitochondrial content, but dysregulated mitochondrial morphology. These changes were accompanied by increased expression of basigin. In the context of COVID-19, CD147/basigin has mainly been discussed to interact with SARS-CoV-2 and to serve as an additional infection route but not as marker for metabolically impaired immune cells (Xia and Dubrovska, 2020). This is surprising as basigin is a metabolic hub and is central for the regulation of cell metabolism. It is highly expressed on a variety of cells, tightly associates with the lactate transporters monocarboxylate transporters (MCTs) and is essential for their cell surface translocation and activities. Basigin also associates with other metabolic proteins including GLUT1 and CD98 (Hahn et al., 2015, Muramatsu, 2016). Here we describe for the first time a correlation between high ROS and basigin levels which suggests basigin as marker for a metabolically dysregulated T cell phenotype in COVID-19 patients. The analysis of established activation markers on T cells and myeloid cells did not reveal any significant changes in COVID-19 patients and we observed only a trend towards increased expression of the T cell activation and exhaustion markers CD25 and PD-1.

The role of basigin in SARS-Cov-2 entry is possibly based on its function as receptor for cyclophilins (CypA), which bind to SARS-CoV proteins thereby allowing virus entry (Chen et al., 2005, Pawlotsky, 2020). In line, CypA inhibitors such as cyclosporine A (CsA) can block virus replication (Tanaka et al., 2013). Even though we detected high CypA levels, especially in critically ill patients, we found no significant traces of SARS-CoV-2 RNA in monocytes or T cells from COVID-19 patients. These data however do not exclude SARS-Cov-2 entry in immune cells *in vitro* as it has been shown by Codo et al (Codo et al., 2020) or in other cell types such as endothelial cells.

The same authors showed elevated mitochondrial ROS and HIF-1a expression in monocytes from COVID-19 patients. Monocyte HIF-1a expression was also increased in our study and we found a hypoxia-related gene set enrichment in monocytes but not T cells. Nevertheless, we detected highly increased cytosolic ROS levels and a ROS-related gene set enrichment in T cells of critically ill patients. But what is the cause of oxidative stress in T cells? COVID-19 patients can suffer from extensive hypoxia (Huang et al., 2020) and COVID-19 pathogenesis is most likely related to hypoxia which can lead to superoxide radical and peroxide generation by mitochondria. In line, COVID-19 T cells showed an increased mitochondrial content but a disturbed morphology with elongated mitochondria and disrupted cristae indicating impaired mitochondrial function, probably leading to ROS production. Mitochondrial impairment and the associated generation of ROS have been shown to stabilize CD147 expression (De Saedeleer et al., 2014), which might explain the strong correlation between ROS levels and basigin expression in T cells. Notably, the upregulation of basigin membrane levels in T cells was not reflected by our RNAseq analyses which is in line with data from by DeSaedeleer et al showing that ROS stabilizes basigin posttranslationally (De Saedeleer et al., 2014).

High basigin levels may lead to suppressed T cell function and proliferation as basigin deficient mice are more active in mixed lymphocyte reaction (Hahn et al., 2015). Furthermore, the related high ROS levels could induce cell death (Hildeman et al., 2003) possibly explaining the reduced T cell numbers in COVID-19 patients which were described here and by others (Lucas et al., 2020, Qin et al., 2020). Severely and critically ill patients were also characterized by low NK cell numbers and increased numbers of B cell blasts. However, our data indicate that immune cell reactivity and metabolism rather than frequency might dictate the outcome of inflammation in COVID-19 patients. In contrast to T cell lymphopenia, the immuno-metabolic dysregulation with high ROS and basigin levels was reversed in the early recovery phase, where severely and critically ill COVID-19 patients improve and are no longer ventilated, indicating that hypoxia may be one important underlying factor for disease outcome. Interestingly, T cell frequencies were significantly lower in diabetic and/or obese patients and T cells and myeloid cells of these patients exhibited decreased glucose uptake. In contrast, cardiovascular disease or age had no impact on T cell numbers and glucose uptake, suggesting an interplay between systemic and T cell metabolism in patients with metabolic disorders. Given the high levels of systemic inflammation the findings of low glucose and fatty acid uptake by T cells and myeloid cells were unexpected, especially as hypoxia can promote fatty acid uptake (Zhang et al., 2017). Increased fatty acid uptake was only found in T cells from SARS-CoV-2 infected patients with none or mild symptoms compared to critically ill patients. Interestingly, T cell fatty uptake capacity did not decrease until four weeks after the onset of symptoms, thus reaching beyond the time point of infection resolution. We speculate that disturbed mitochondrial function in severely ill patients may prevent fatty acid degradation, thereby reducing fatty acid uptake.

Our data support the hypothesis that COVID-19 pathogenesis is related to hypoxia and oxidative stress. Besides ROS in lymphocytes, neutrophils of critically ill patients with COVID-19 may produce ROS to drive pathological host responses as it has been recently proposed by Laforge and colleagues (Laforge et al., 2020). They suggested that ROS-induced tissue damage, thrombosis and red blood cell dysfunction could be targeted with free radical scavengers. Such an approach could also ameliorate T cell function and is currently tested in an ongoing study treating COVID-19 patients with antioxidant drugs such as vitamin C (Cavezzi et al., 2020).

Importantly, the clinical use of the glucocorticoid dexamethasone is beneficial for COVID-19 patients (Group et al., 2020) but its mechanisms of action are not yet fully understood and its effects on immune cells are broad. Besides its widely established role as immunosuppressant, dexamethasone can support T cell viability (Franchimont et al., 2002). Surprisingly, in our experiments, treatment with dexamethasone was effective in preventing intracellular ROS accumulation without affecting activation and cytokine production in expanded T cells of COVID-19 patients. We therefore propose that the effects of dexamethasone in COVID-19 patients are at least partially due to an immunometabolic modulation. Future studies should address if other agents specifically targeting T cell metabolism could improve the outcomes of critically ill COVID-19 patients.

Another treatment option might be a direct targeting of basigin. Meplazumab, a humanized anti-CD147 antibody, inhibited SARC-CoV-2 replication (Wang et al., 2020). According to the preliminary report on the clinical trial study NCT04275245, meplazumab improved the outcome of patients with COVID-19-associated pneumonia with a favorable safety profile (Bian et al., 2020). Importantly, basigin blockade does not only target the virus replication but also attenuates T cell chemotaxis induced by CyPA (Lv et al., 2018) and can therefore inhibit COVID-19-associated inflammation. CyPA itself could also be a promising target structure as increased CyPA levels were detected in the severe/critical COVID-19 patients that decreased in the convalescence. Thus, CyPA might also be involved in dysregulated inflammation in COVID-19 patients. Replication of coronaviruses and several other viruses requires cyclophilins which can be targeted by the established immunosuppressant cyclosporin A (CsA), which, in addition to targeting calcineurin, blocks cyclophilin (Tanaka et al., 2013). Furthermore, Alisporivir, a non-immunosuppressive derivative of CsA also limits the replication of human coronavirus HCoV-NL63 (Carbajo-Lozoya et al., 2014).

We propose here that hypoxia and ROS induce immune cell basigin expression, contributing to metabolic dysfunction and a dysregulation of inflammation in COVID-19 patients. Even though future mechanistic studies will test this hypothesis, cellular basigin expression and cyclophilin blood levels might serve as both prognostic parameters and therapeutic targets in COVID-19. Furthermore, our study strengthens the rationale for the use of agents that reduce oxidative stress in COVID-19 therapy.

## Data Availability

Data are not available on external repositories

## Declarations

## Acknowledgements

We thank Marcus Kielmanowicz, Monika Wehrstein und Rüdiger Eder for technical assistance. We thank the members of the RCI FACS Core Facility for expert technical assistance. RNA-sequencing was conducted at the NGS Core of the Regensburg Center for Interventional Immunology (RCI, University Regensburg and University Medical Center Regensburg, Germany).The authors declare no competing interests.

This work was supported by the Bavarian Ministry of Science and Arts. PJS is supported by Else Kröner Fresenius Foundation. PH is supported by the Bavarian Ministry of Science and Arts.

## Author contributions

PJS, KR conceived the project, performed experiments, analyzed data and wrote the manuscript, MK conceived the project and wrote the manuscript, KS analyzed data and supervised experiments, JK, NK, SMD, KK performed experiments and analyzed data, CB, CM, AP, GS, JR performed experiments, DL, BG, FG, ML, MM, PHa, CB, RB, AG, BS, FHa, FHi, WH provided critical resources, DH, FL, TP, DW, HP provided critical resources and supervised experiments, CB performed EM experiments, PHo performed cell sorting and MR transcriptome analyses.

## ORCID

P.J.S: 0000-0002-1521-6213

M.R.: 0000-0003-3992-932X

D.H.: 0000-0002-8790-4584

F.L.: 0000-0001-5252-287X

K.R.: 0000-0002-7116-1368

F.G.: 0000-0002-0952-0841

P.Ha.: 0000-0003-3894-5053

## Methods

### Study participants

This study involved 61 subjects consisting of 45 patients with confirmed SARS-CoV-2 infection, detected in a nasopharyngeal swab or a respiratory sample using routinely established RT-PCR (**Table 1**) and 16 uninfected controls. The study was approved by the appropriate Institutional Review Board (University Hospital Regensburg, No. 20-1785-101) and conformed to the principles outlined in the Declaration of Helsinki. Included were all patients with positive SARS-CoV-2 testing, excluded was one patient with first-diagnosis of leukemia. Controls were asymptomatic healthy individuals with age median of 46.7 (IQR 37.25-46.7) and male sex in 47%. Disease severity was assessed as described previously (Song et al., 2020, Hadjadj et al., 2020). Briefly, none/mild cases presented with no or mild symptoms and no pulmonary infiltrate; moderate cases presented with respiratory symptoms such as cough and/or shortness of breath and a pulmonary infiltrate; severe/critical cases included patients with persistent tachypnea >30/min, blood oxygen saturation <93%, rapid progress of pulmonary infiltrates within 24 hours, respiratory failure or requirement for mechanical ventilation. Clinical, laboratory, treatment and outcome data were extracted from electronic medical records using standard procedures and confidentiality measures. Routine testing included a differential blood count and biochemical tests. Antibody response was defined by presence of SARS-CoV-2 specific IgG antibodies in serum.

### Sample collection and processing

All whole-blood samples analyzed for immune cell frequencies and single cell metabolic assays were analyzed freshly without cryopreservation. Blood was collected using Lithium-Heparin-Tubes (Sarstedt, Nümbrecht, Germany). For ACK lysis, samples were divided in 1 ml portions and incubated for 5 min in 20 ml ACK buffer 1X (6X: 0.155M NH4Cl, 0.01M KHCO3 and 0.1 mM EDTA), afterwards 30 ml of PBS were added, centrifuged (5 min, 523G) and supernatant was discarded. Lysis was repeated using 10 ml ACK and 20 ml PBS per sample. If erythrocytes were still visible in the pellet, a third lysis step (5 ml ACK and 20 ml PBS) was performed and finally cells were washed in 20 ml PBS. Finally, cells were counted in 5 ml PBS + 2 % FCS using the CASY Cell Counter system (Cursor Setting: CL 6.50 μM, NL 4.50 μM, CR 30 μM, NR 30 μM). For *in vitro* culture assays, blood samples were processed using the Ficoll gradient centrifugation and mononuclear cells were cryopreserved until analysis. After thawing, cells were cultured in complete RPMI medium (Gibco), supplemented 2-mercaptoethanol (Gibco), with or without the addition of anti-CD3/anti-CD28 coated beads (Gibco), with or without 100 nM or 500nM Dexamethasone (Jenapharm) which was dissolved in medium.

### Determination of immune cell populations

For analysis of surface marker expression, 0,2-1×10^6^ cells/tube were stained with respective surface markers (see list of antibodies below). For determination of regulatory T cells intracellular FOXP3 expression was measured using the Intracellular Fixation and Permeabilization Buffer Set according to manufacturer’s instructions (ThermoFisher). In brief, after surface marker staining (anti-CD3, anti-CD4), intracellular unspecific binding sites were blocked by incubation with 2 % rat serum for 15 minutes and subsequently, intracellular staining (anti-FOXP3) was performed.

### Single cell metabolic assays

Intracellular reactive oxygen species (ROS) were determined after surface marker staining (anti-CD3, anti-CD11b, anti-CD14, anti-CD66b), using 10 μM 2’,7’-dichlorofluorescin diacetate (Sigma, D6883) in a cell culture incubator 37° for 20 minutes in closed tubes. Cells were washed once with 3 ml cold PBS and measured immediately. Mitochondrial content was assessed by staining with Mitotracker Green FM. Cells were resuspended in RPMI 1640 supplemented with 2 mM L-Glutamine and treated with 15 nM Mitotracker and Cyclosporine A for 1 hour under cell culture conditions. Afterwards, surface staining was performed. Glucose and fatty acid uptake was measured using the fluorescent glucose analog 2-[N-(7-nitrobenz-2-oxa-1,3-diazol-4-yl)amino]-2-deoxy-D-glucose (2-NBDG) and BODIPY 500/510 C1, C12 dye (both Thermo Fisher). Cells were stained with surface antibodies as described above and incubated in with 2-NBDG (45 min) or BODIPY 500/510 C1, C12 (20 min). Fluorescence of selected immune cell subpopulations was measured by flow cytometry.

### Electron microscopical analyses

For transmission electron microscopic analyses, cell pellets were fixed in Karnovsky-fixative and enclosed with 4% low-melting-agarose (Invitrogen, Germany). For the embedding process including post-fixation with osmium tetroxide, dehydration, infiltration with EPON (all reagents from EMS, Science Services, Germany) the LYNX microscopy tissue processor (Reichert-Jung, Wetzlar, Germany) was used. Semi-thin-sections (75μm), for the selection of relevant areas, and ultra-thin sections (80nm) for the ultrastructural analyses were cut by use of the Reichert Ultracut S Microtome (Leica-Reichert, Wetzlar, Germany). Ultra-thin-sections were contrasted with aqueous 2%-uranyl-acetate- and 2%-lead-citrate solution for 10 minutes each. The Electron-microscopic analysis was performed using the LEO 912AB electron-microscope (Zeiss, Oberkochen, Germany).

### Cell sorting and RNA-seq library preparation

For RNASeq experiments, cryopreserved PBMC were thawed and stained with anti-CD3, anti-CD4, anti-CD8 and anti-CD14. Live, single cells with leukocyte FSC/SSc characteristics were sorted into CD14+CD3-monocytes, CD14-CD3+CD4+CD8-CD4 T cells and CD14-CD3+CD4-CD8+ CD8 T cells by four-way sorting on a BD FACSAria IIu (Becton Dickinson, Heidelberg, Germany). Sorted cells were centrifuged and further processed for RNASeq immediately. Total cellular RNA was isolated from sorted blood cells using the RNeasy Micro Kit (Qiagen) according with manufacturer’s instructions. RNA was quantified using the NanoDrop (peqLab) and the quality was assessed using the RNA ScreenTape Kit (Agilent). Generation of dsDNA libraries for Illumina sequencing was carried out using the SMART-Seq Stranded Kit (Takara Bio) according to manufacturer’s instructions. The quality of dsDNA libraries was checked with the High Sensitivity D1000 ScreenTape Kit (Agilent) and concentrations were determined with the Qubit dsDNA HS Kit (Thermo Fisher Scientific). Sequencing was performed using the Illumina NextSeq550 sequencer.

### RNA-seq analysis

Sequencing reads were mapped to human (GRCh38) or viral (SARS-CoV-2, NC_045512.2) genome using STAR v2.5.3a (Dobin et al., 2013). SARS-CoV-2 infection of A549 cells was performed and published by Blanco-Melo et al (Blanco-Melo et al., 2020). The human GRCh38 genome index incorporated gene annotation from GENCODE 44 (release 27) to aid in spliced alignment. Tables of raw uniquely mapped read counts per human gene were generated during mapping using the built-in --quantMode GeneCounts option in STAR. Differential expression analysis was carried out on raw gene counts using edgeR 3.20.8 (Robinson et al., 2009) in R (3.4.3). Pairwise comparisons of indicated data sets were done using the quasi-likelihood test. Heatmaps of differentially expressed genes shown in Figure 5 used batch-corrected, normalized and scaled CPM (counts per million) data and were generated using the heatmap.2 function of the gplots package in R. Volcano plots were generated using the ggplot2 (v3.1.0) package in R. The rank-based gene set enrichment tests in Figure 5 were done using the fry function of the limma package (Ritchie et al., 2015) and plotted using the barcodeplot function in R. Gene-sets were defined in the hallmark gene set collection (Liberzon et al., 2015) and basigin interaction partners were defined by STRING (Szklarczyk et al., 2019).

### Cyclophilin A plasma levels

Cyclophilin A levels were determined in plasma samples by a commercially available enzyme-linked immunosorbent assay (ELISA, Sigma-Aldrich) according to the manufacturer’s protocol.

### Statistics

GraphPad Prism was used for statistical analyses, using ANOVA with post-hoc Bonferroni test and Mann-Whitney U test. For paired ANOVA analyses with missing values, mixed-effects model with the Geisser-Greenhouse correction was used. Significance levels were *P < 0.05; **P < 0.01; ***P < 0.001. Correlations were computed using the Spearman correlation test. Statistical tests used for clinical parameters: Kruskal-Wallis (age), chi-squared test nach Pearson (all categorical variables), students t-test (ferritin), one-way ANOVA (leukocytes and all other laboratory values).

List of reagents.

List of reagents
| Antibody | Source | Identifier |
| --- | --- | --- |
| Anti-Human CD3 (clone SK7) | Invitrogen (Thermo Fisher Scientific; Waltham, MA, US) | Cat# 45-0036-42 |
| Anti-Human CD3 (clone SK7) | BD Biosciences (Franklin Lakes, NJ, US) | Cat# 341111/ 345763 |
| Anti-Human CD3 (clone SK7) | BioLegend (San Diego, CA, US) | Cat# 344828 |
| Anti-Human CD3 (clone SK7) | BD Biosciences | Cat# 345763 |
| Anti-Human CD4 (clone SK3) | BD Biosciences | Cat# 345770 |
| Anti-Human CD4 (clone RPA-T4) | BD Biosciences | Cat# 560345 |
| Anti-Human CD8 (clone SK1) | BioLegend | Cat# 344712 |
| Anti-Human CD8 (clone SK1) | BD Biosciences | Cat# 560179 |
| Anti-Human CD11b (clone M1/70) | BioLegend | Cat# 101230 |
| Anti-Human CD14 (clone 63D3) | BioLegend | Cat# 367124 |
| Anti-Human CD16 (clone 3G8) | BD Biosciences | Cat# 555406 |
| Anti-Human CD19 (clone SJ25C1) | BD Biosciences | Cat# 345789 |
| Anti-Human CD20 (clone L27) | BD Biosciences | Cat# 345792 |
| Anti-Human CD25 (clone 2A3) | BD Biosciences | Cat# 341011 |
| Anti-Human CD33 (clone P67.6) | BioLegend | Cat# 366608 |
| Anti-Human CD38 (clone HB7) | BD Biosciences | Cat# 335825 |
| Anti-Human CD45RA (clone HI100) | BD Biosciences | Cat# 563031 |
| Anti-Human CD56 (clone HCD56) | BioLegend | Cat# 318328 |
| Anti-Human CD66b (clone G10F5) | BioLegend | Cat# 305112 |
| Anti-Human CD98 (clone REA387) | Miltenyi Biotec | Cat# 130-105-664 |
| Anti-Human CD147 (clone HIM6) | BioLegend | Cat# 306214 |
| Anti-Human CD197 (clone G043H7) | BioLegend | Cat# 353207 |
| Anti-Human CD279 (clone EH12.1) | BD Biosciences | Cat# 563076 |
| Anti-Human HLA-DR (clone G46-6) | BD Pharmigen | Cat# 561358 |
| Anti-Human FOXP3 (clone PCH101) | eBioscience (Thermo Fisher Scientific) | Cat# 12-4776-42 |
| Anti-Human IgA (clone IS11-8E10) | Miltenyi Biotec | Cat# 130-114-004 |
| Anti-Human IgG (clone IS11-3B2.2.3) | Miltenyi Biotec | Cat# 130-119-962 |

| Anti-Human IgM (clone MHM-88) | BioLegend | Cat# 314520 |
| --- | --- | --- |
| Reagent | Source | Identifier |
| Ammonium Chloride | Merck (Darmstadt, Germany) | Cat# 101145 |
| 2',7'-Dichlorofluorescein diacetate | Sigma-Aldrich (St. Louis, MO, US) | Cat# D6883 |
| 2-[N-(7-nitrobenz-2-oxa-1,3-diazol-4-yl)amino]-2-deoxy-D-glucose (2-NBDG) | Thermo Fisher Scientific | Cat# N13195 |
| Brilliant Stain Buffer | BD Biosciences | Cat# 563794 |
| BODIPY 500/510 C1, C12 | Thermo Fisher Scientific | Cat# D3823 |
| Cyclosporine A (Sandimmune® 50 mg/ml) | Novartis (Basel, Schweiz) |  |
| Dulbeccos Phosphate Buffered Saline (DPBS) | Gibco (Thermo Fisher Scientific) | Cat# 14190-094 |
| Ethylendiamine-tetraacetic-acid disodium salt dihydrate | Sigma-Aldrich | Cat# E5134 |
| Fetal Calf Serum | Sigma-Aldrich | Cat# F7524-500 ML |
| Fixable Viability Dye eF780 | eBioscience (Thermo Fisher Scientific) | Cat# 65-0865-14 |
| Intracellular Fixation and Permeabilization Buffer Set | eBioscience (Thermo Fisher Scientific) | Cat# 88-8824-00 |
| L-Glutamine | PAN Biotech (Aidenbach, Germany) | Cat# P04-80050 |
| Mitotracker Green FM | Invitrogen (Thermo Fisher Scientific) | Cat# M7514 |
| RPMI 1640 | Gibco (Thermo Fisher Scientific) | Cat# 31870-025 |
| Dexamethasone | Jenapharm, mibe GmbH |  |
| 2-Mercaptoethanol | Gibco | Cat# 31350010 |
| Cyclophilin A ELISA | Sigma-Aldrich | Cat#RAB1402 |
| Devices | Source |  |
| Megafuge 3.0R | Haereus (Hanau, Germany) |  |
| CASY Cell Counter | OMNI Life Science (Bremen, Germany) |  |
| BD FACSCelesta Flow Cytometer | BD Biosciences |  |
| Software | Source | Identifier |
| FlowJo (V 9.9.6, 10.6.2, 10.7.1) | Tree Star Inc (BD Life sciences) | https://www.flowjo.com |
| Prism | GraphPad |  |
| edgeR 3.20.8 |  | Robinson et al., 2009 |

---

**Figure S1.**
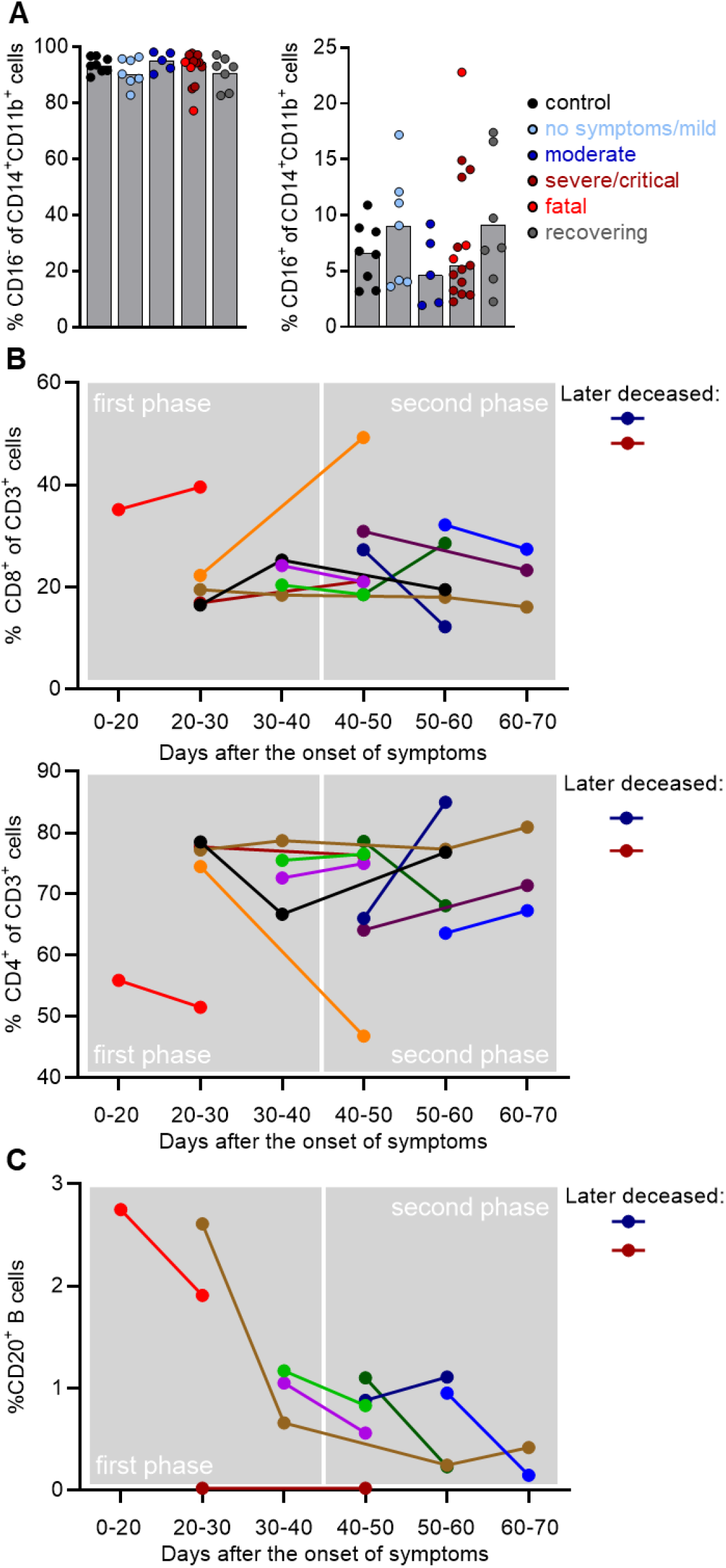
Maintained monocyte but altered T cell subpopulation distribution in COVID-19 patients. Blood of donors was drawn and processed on the same day. Erythrocytes were removed by ACK lysis and immune cell populations were determined by specific surface marker staining and subsequent analysis by flow cytometry. (**A**) Percentage of CD16- and CD16+ cells among CD14+CD11b+ monocytes was determined. Shown is the median, each symbol represents one donor. Frequencies of CD8+ and CD4+ T cell subpopulations (**B**) and CD20+ B cells (**C**) are shown in paired patient samples over time and clustered to groups determined by the number of days after the onset of symptoms.

**Figure S2.**
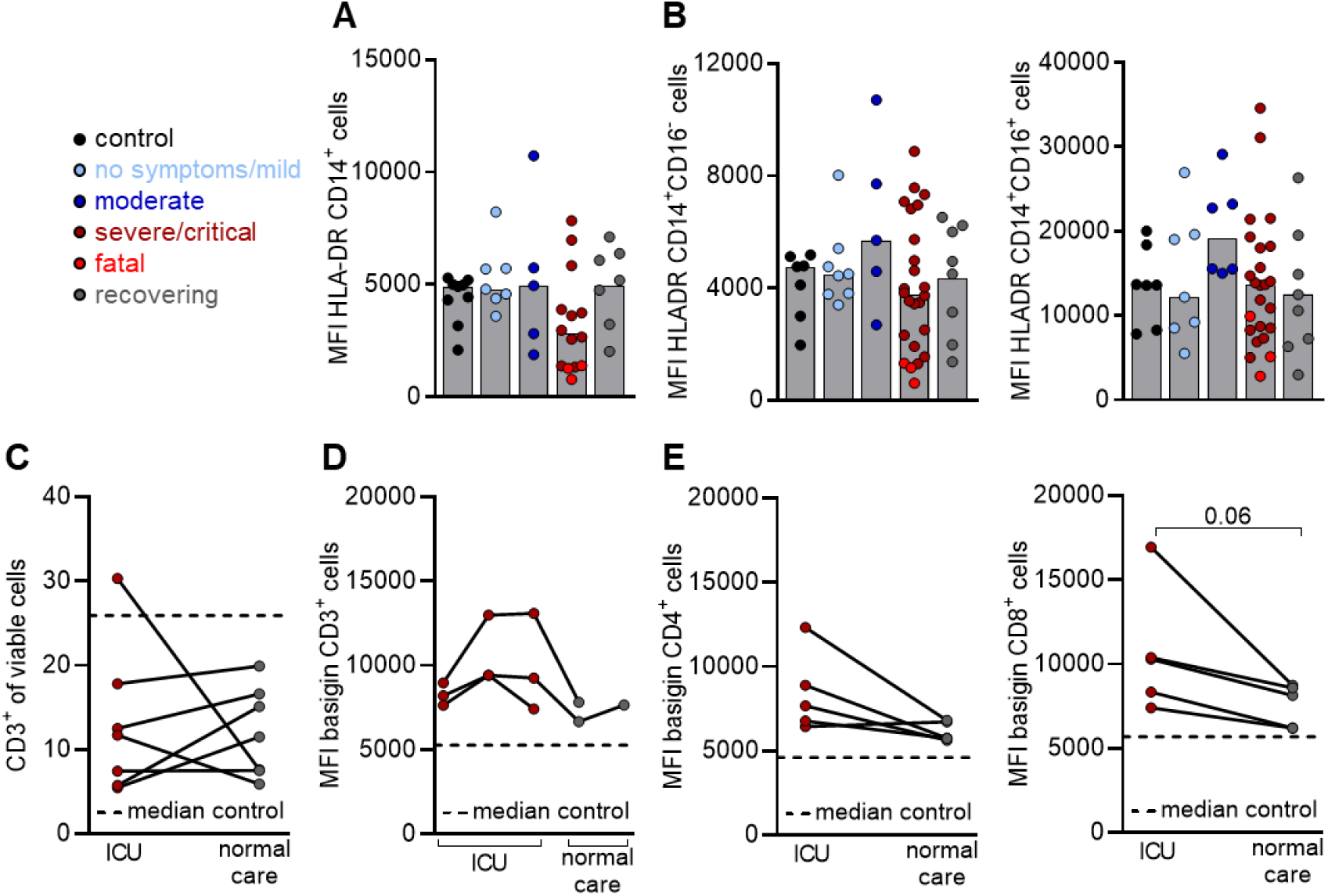
Preserved HLA-DR expression on monocytes but increased basigin expression, reduced during recovery, on T cells of progressed COVID-19 patients. Blood of donors was drawn and processed on the same day. Erythrocytes were removed by ACK lysis and immune cell populations were stained for population-specific surface markers. HLA-DR expression as an indicator for MHC-II surface expression was analyzed on CD14+ monocytes (**A**) and the respective CD16- and CD16+ (**B**) subpopulations. Shown is the median, each symbol represents one donor. Frequencies of CD3+ T cells (**C**) and basigin expression on CD3+ T cells (**D**) and CD4+ and CD8+ T cell subpopulations (**E**) were analyzed in paired patient samples taken at the intensive care unit (ICU) and after returning to normal care unit. Shown are paired patient samples (paired Student’s t-test).MF FA uptake CDS-T cel 5 MF FA **uptake** CD4-**T cells Q MFI FA uptake CD4~ cells** μ **MFI CD98 of CD3* cells**

**Figure S3.**
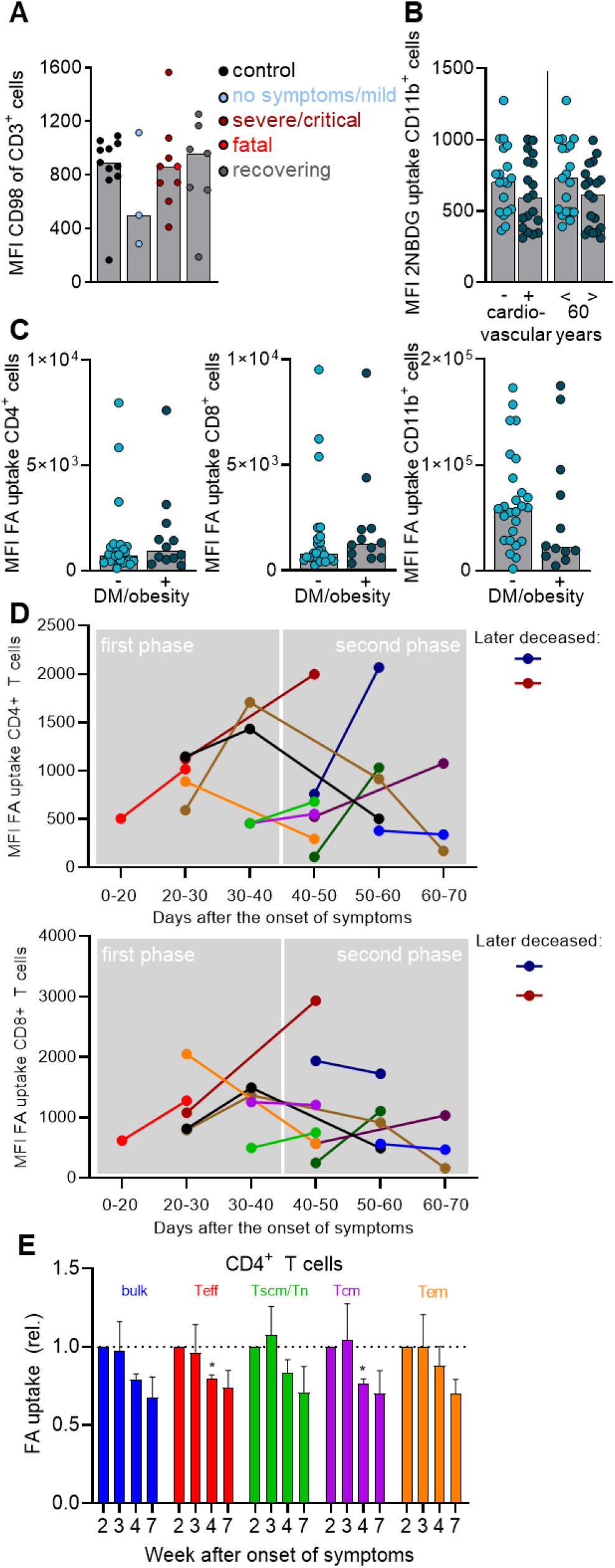
CD98 expression, glucose and fatty acid uptake by immune cells of COVID-19 patients. Blood of donors was drawn and processed on the same day. Erythrocytes were removed by ACK lysis and expression of surface molecules or nutrient uptake of immune cell populations was determined by flow cytometry. (**A**) CD98 surface expression was analyzed in all groups. Shown is the median, each symbol represents one donor. (**B**) Comparison of 2NBDG uptake in CD11b+ myeloid cells of patients in relation to cardiovascular disease or age. Shown is the median, each symbol represents one donor. (**C**) BODIPY 500/510 C1, C12 uptake as a measure of fatty acid (FA) uptake in CD4+, CD8+ T cells and CD11b+ myeloid cells of diabetic and/or obese patients versus non-diabetic and/or obese patients. Shown is the median, each symbol represents one donor. (**D**) FA uptake in CD4+ and CD8+ T cells of paired patient samples over time, with clustering of samples into groups according the number of days after the onset of symptoms. (**E**) Analysis of FA uptake in CD4+ T cells from 3 patients of one family with mild COVID-19 over time. Shown is the mean and SEM of values normalized to the first blood sample drawn (Mixed-effects model with the Geisser-Greenhouse correction, with Bonferroni’s multiple comparisons test, *p<0.05).

**Figure S4.**
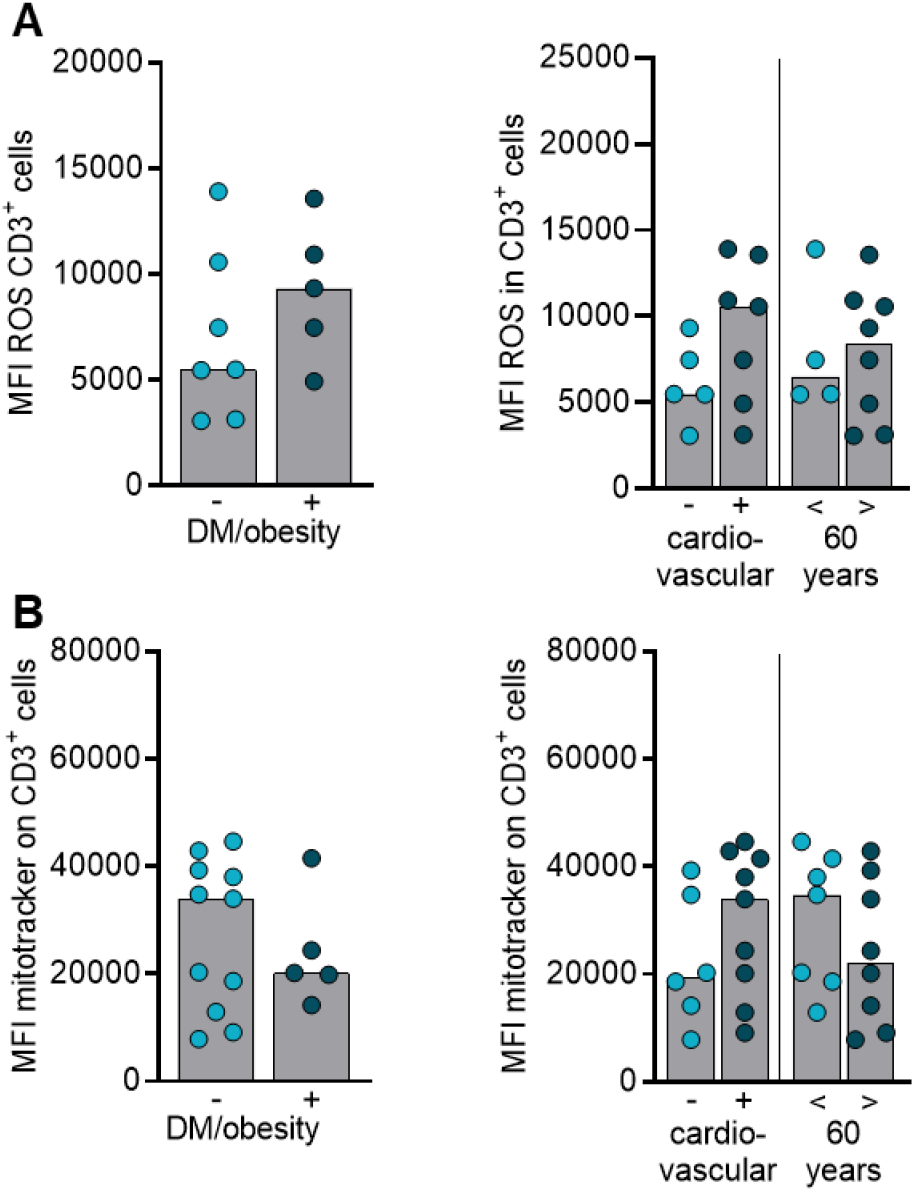
Increased ROS levels and mitochondrial content are not related to diabetes and/or obesity, cardiovascular disease or age. Blood of donors was drawn and processed on the same day. Erythrocytes were removed by ACK lysis and immune cell populations were stained for population-specific surface markers. (**A**) Cytosolic ROS level in CD3+ T cells, determined by DCFDA staining and analyzed by flow cytometry, was correlated to pre-existing diabetes and/or obesity, cardiovascular disease or age. (**B**) Mitochondrial content in CD3+ T cells, determined by mitotracker staining and analyzed by flow cytometry, in relation to diabetes and/or obesity, cardiovascular disease or age.

**Figure S5.**
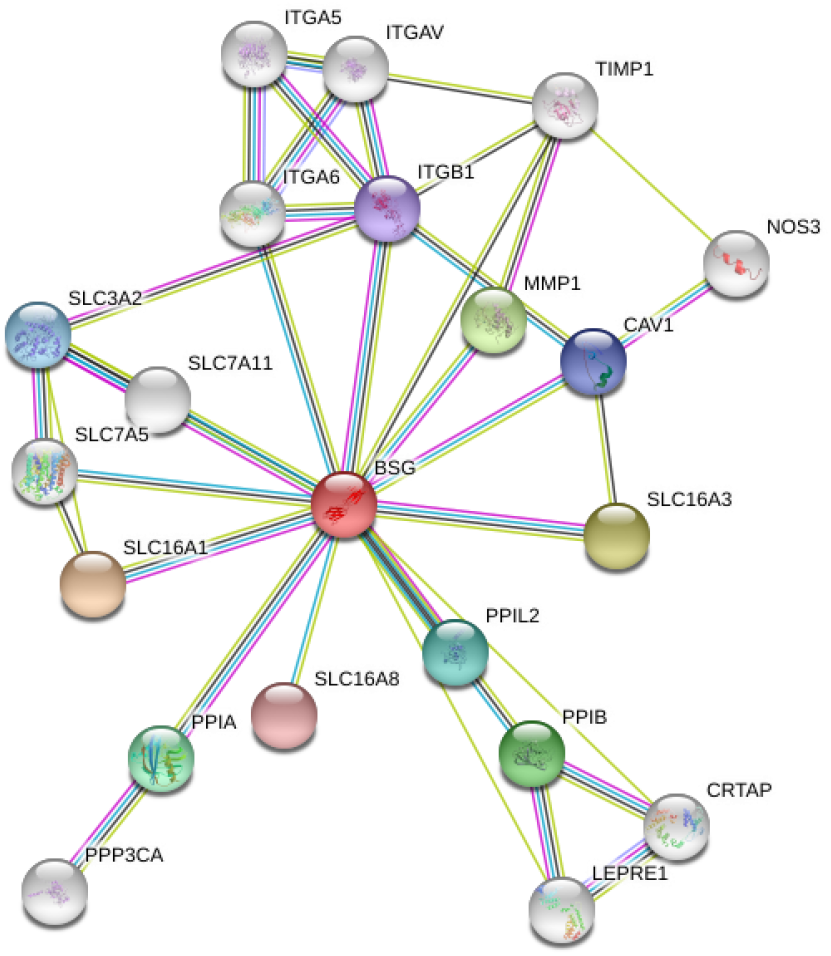
Interaction partners of basigin. Basigin interaction partners were defined by STRING (Szklarczyk et al., 2019).

**Figure S6.**
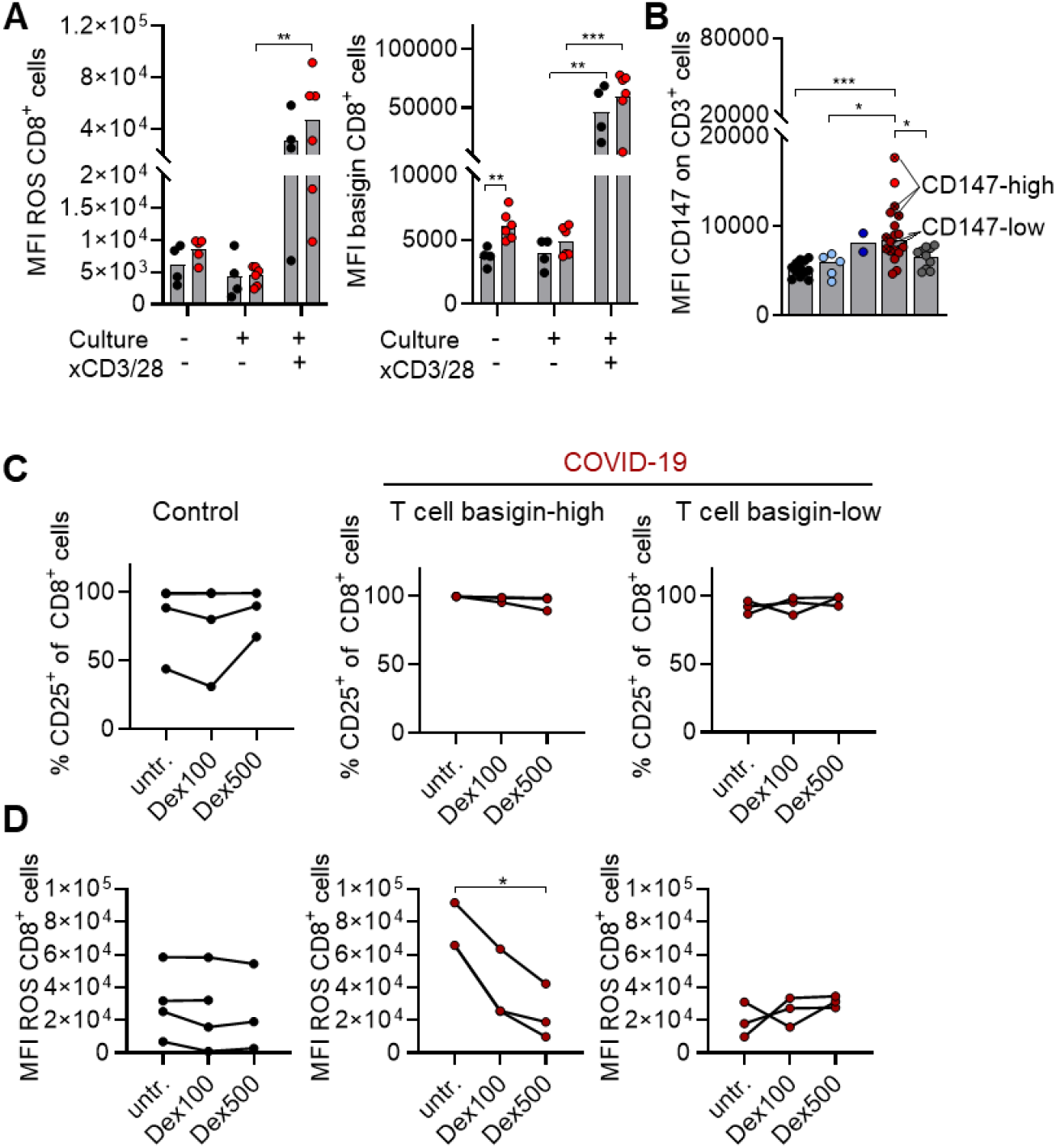
Activation induced ROS accumulation in COVID-19 T cells is reversible through dexamethasone treatment. Mononuclear cells were isolated from controls or COVID-19 patients from the severe/critical group and cryopreserved until analysis. (**A**) ROS and basigin levels were determined in CD8+ T cells of controls and severe/critically ill patients before and after a 6 days culture in medium containing 2-mercaptoethanol with or without anti-CD3/anti-CD28 stimulation. Shown are median levels and a single symbol for each donor (paired Student’s t-test, *p<0.05, **p<0.01, ***p<0.001). (**B**) Basal basigin expression on CD3+ T cells, also depicted as **Figure 2D**. (**C,D**) Mononuclear cells of controls and COVID-19 patients with high and low basal basigin expression, as pre-defined in (**B**) were anti-CD3/anti-CD28 stimulated for six days in the presence or absence of increasing concentrations of dexamethasone (100 nM, 500 nM) and percentage of CD25+CD8+ T cells (**C**) and cytosolic ROS levels, determined by DCFDA staining (**D**), were assessed. Shown are paired samples from each donor, represented by a symbol (one-way ANOVA, Bonferroni multiple comparisons test, *p<0.05, **p<0.01).

## Notes

### Competing Interest Statement

The authors have declared no competing interest.

### Funding Statement

This work was supported by the Bavarian Ministry of Science and Arts. PJS is supported by Else Kroener Fresenius Foundation. PH is supported by the Bavarian Ministry of Science and Arts.

### Author Declarations

The study was approved by the appropriate Institutional Review Board (University Hospital Regensburg, No. 20-1785-101) and conformed to the principles outlined in the Declaration of Helsinki.

### Summary of Updates

Author affiliations updated

